# Artemether-lumefantrine with or without single-dose primaquine and sulfadoxine-pyrimethamine plus amodiaquine with or without single-dose tafenoquine to reduce *Plasmodium falciparum* transmission: a phase 2 single-blind randomised clinical trial in Ouelessebougou, Mali

**DOI:** 10.1101/2024.02.27.24303370

**Authors:** Almahamoudou Mahamar, Merel Smit, Koualy Sanogo, Youssouf Sinaba, Sidi M. Niambele, Adama Sacko, Oumar M Dicko, Makonon Diallo, Seydina O Maguiraga, Yaya Sankaré, Sekouba Keita, Siaka Samake, Adama Dembele, Kjerstin Lanke, Rob ter Heine, John Bradley, Yahia Dicko, Sekou F. Traore, Chris Drakeley, Alassane Dicko, Teun Bousema, Will Stone

**Affiliations:** Malaria Research and Training Centre, Faculty of Pharmacy and Faculty of Medicine and Dentistry, University of Sciences Techniques and Technologies of Bamako, Bamako, Mali; Department of Medical Microbiology and Radboud Center for Infectious Diseases, Radboud University Medical Center, Nijmegen, the Netherlands; Department of Pharmacy and Radboud Center for Infectious Diseases, Radboud University Medical Center, University of Nijmegen, Nijmegen, the Netherlands; MRC International Statistics and Epidemiology Group, London School of Hygiene and Tropical Medicine, London, UK; Department of Infection Biology, London School of Hygiene & Tropical Medicine, London, UK, WC1E7HT

**Author notes:** Authors contributed equally (Almahamoudou Mahamar, Merel Smit, Chris Drakeley, Alassane Dicko, Teun Bousema, Will Stone). **Corresponding author:** Almahamoudou Mahamar. **Alternate corresponding author:** Alassane Dicko.

**Keywords:** Gametocytes, Artemether-lumefantrine, Primaquine, Sulfadoxine-pyrimethamine plus amodiaquine, Tafenoquine, Transmission-blocking

## Abstract

**Background:** Artemether-lumefantrine is widely used for uncomplicated *Plasmodium falciparum* malaria; sulfadoxine-pyrimethamine plus amodiaquine is used for seasonal malaria chemoprevention. We determined the efficacy of artemether-lumefantrine with and without primaquine and sulfadoxine-pyrimethamine plus amodiaquine with and without tafenoquine for reducing gametocyte carriage and transmission to mosquitoes.

**Methods:** In this phase 2, single-blind, randomised clinical trial conducted, asymptomatic individuals aged 10-50 years with *P. falciparum* gametocytaemia were randomised (1:1:1:1) to receive either artemether-lumefantrine, artemether-lumefantrine with a single dose of 0·25 mg/kg primaquine, sulfadoxine-pyrimethamine plus amodiaquine or sulfadoxine-pyrimethamine plus amodiaquine with a single dose of 1·66 mg/kg tafenoquine. All trial staff other than the pharmacist were blinded. The primary outcome was the median within person percent change in mosquito infection rate in infectious individuals from baseline to day 2 (artemether-lumefantrine groups) or 7 (sulfadoxine-pyrimethamine plus amodiaquine groups) post treatment, assessed by direct membrane feeding assay. This study is registered withClinicalTrials.gov, NCT05081089.

**Findings:** Between 13 Oct and 16 Dec 2021, 1290 individuals were screened and 80 were enrolled and randomly assigned to one of the four treatment groups (20 per group). In individuals who were infectious before treatment, the median percentage reduction in mosquito infection rate 2 days after treatment was 100% (IQR 97·2-100; n=19, p=0·026) with artemether-lumefantrine and 100% (100-100; n=19, p=0·0001) with artemether-lumefantrine with primaquine. Only two individuals infected mosquitoes on day 2 after artemether-lumefantrine and none at day 5. In contrast, the median percentage reduction in mosquito infection rate 7 days after treatment was 63·60% (IQR 0·62 to 100, n=20, p=0·009) with sulfadoxine-pyrimethamine plus amodiaquine and 100% (100-100; n=19, p<0·0001) with sulfadoxine-pyrimethamine plus amodiaquine with tafenoquine.

**Interpretation:** These data support the effectiveness of artemether-lumefantrine alone for preventing nearly all mosquito infections. In contrast, there was considerable post-treatment transmission after sulfadoxine-pyrimethamine plus amodiaquine where the addition of a transmission-blocking drug may be beneficial in maximizing its community impact.

**Funding:** Bill & Melinda Gates Foundation

**Brief summary:** Sulfadoxine-pyrimethamine plus amodiaquine is commonly used for seasonal malaria chemoprevention. Artemether-lumefantrine is the most widely used treatment regimen for uncomplicated *Plasmodium falciparum* malaria, but studies to date have shown inconsistent activity of artemether-lumefantrine against *P. falciparum* gametocytes. This study shows considerable post-treatment transmission after sulfadoxine-pyrimethamine plus amodiaquine but near complete prevention of mosquito infection after artemether-lumefantrine, even without primaquine. The addition of 8-aminoquinolines reduced transmission with both combinations.

## Introduction

Artemisinin-based combination therapies (ACT) retain excellent efficacy for treatment of uncomplicated malaria on the African continent, despite concerning evidence for reduced sensitivity of parasites in East Africa for artemisinins.^1^ ACTs rapidly clear asexual parasites but have variable activity against mature gametocytes that are the parasite life stage that can be transmitted to mosquitoes. A pooled analysis of post treatment microscopy data indicated that artemether-lumefantrine may be the most potent ACT in terms of gametocyte clearance whilst gametocyte persistence is markedly longer after the drug combination dihydroartemisinin-piperaquine.^2^ Importantly, post-treatment gametocyte carriage is an imperfect approximation of malaria transmission potential. Transmissible gametocytes may persist at submicroscopic densities after treatment and some antimalarial drugs may sterilize gametocytes before these are removed from the circulation.^3,4^ Although artemether-lumefantrine is the most widely used treatment regimen for uncomplicated *Plasmodium falciparum* worldwide,^5^ there are currently no reliable data on malaria transmission post-artemether-lumefantrine. Five studies that performed mosquito feeding assays post-artemether-lumefantrine reached different conclusions. Three studies observed non-negligible transmission between 7 to 14 days after artemether-lumefantrine^6,7^ or artemether-lumefantrine with primaquine^8^ while two others found near complete abrogation of transmission one week after initiation of artemether-lumefantrine treatment without primaquine.^9,10^ The only study that assessed transmission to mosquitoes before and after artemether-lumefantrine treatment saw only one individual (2·0%; 1/49) infect mosquitoes at day 7 after treatment initiation, with mosquito infection rates markedly reduced compared to pre-treatment.^9^ Quantifying post-treatment transmission potential after the most commonly used antimalarial is highly relevant: if there is substantial transmission after treatment with artemether-lumefantrine, the addition of a specific gametocytocide may be considered. The WHO recommends that to reduce onward transmission of *P. falciparum*, ACTs may be combined with a single low-dose of primaquine (0·25 mg/kg),^11^ which has potent but short lived gametocytocidal activity. Although this recommendation is currently limited to areas of low transmission, it has received new relevance in sub-Saharan Africa since the emergence of artemisinin resistance on the African continent.^12^ As a *P. falciparum* gametocytocide, a single low-dose of 0·25 mg/kg primaquine when combined with ACTs causes an immediate and effective reduction in the transmission of parasites^13,14^ and is safe for use without prior glucose-6-phosphate dehydrogenase (G6PD) status testing.^15^ It is currently unclear whether the addition of primaquine to artemether-lumefantrine is beneficial or whether artemether-lumefantrine already sufficiently prevents post-treatment transmission.

In addition, a recent study with tafenoquine, which is being developed as longer-lasting alternative to primaquine for preventing *P. vivax* relapse^16^ demonstrated that a single low-dose of tafenoquine (1·66 mg/kg) in combination with dihydroartemisinin-piperaquine completely prevented infectivity within 7 days after initiation of treatment in Malian adults and children.^17^ Though this trial showed that tafenoquine can prevent transmission when co-administered with dihydroartemisinin-piperaquine, the effect appeared to be slower than primaquine.^18^ Recent observations that Indonesian soldiers given dihydroartemisinin-piperaquine with or without 300 mg tafenoquine showed similar rates of *P. vivax* relapse,^19^ suggest that dihydroartemisinin-piperaquine may negatively impact tafenoquine efficacy. The combination of tafenoquine with non-ACTs to clear *P. falciparum* gametocytes and prevent transmission has not been tested. Sulfadoxine-pyrimethamine plus amodiaquine is a non-artemisinin based combined anti-malarial treatment that is the only antimalarial recommended for systematic mass administration in the form of seasonal malaria chemoprevention (SMC).^20^ Prior studies showed considerable post-treatment transmission potential following sulfadoxine-pyrimethamine plus amodiaquine with infectivity and mosquito infection rate unaffected for the first 7 days post-treatment and reductions in gametocytaemia only observed after 28 days.^7,14^ Increased gametocyte densities in the first 2 weeks following sulfadoxine-pyrimethamine plus amodiaquine will limit any effect of SMC on transmission.^7^ The combination of sulfadoxine-pyrimethamine plus amodiaquine with a single low-dose tafenoquine has never been tested.

In the current study, we assessed the two widely used malaria treatments in combination with single low-dose of gametocytocidal, transmission-blocking drugs in Malian children and adults.

## Methods

### Study design and participants

This four-arm, single-blind, phase 2 randomised controlled trial was conducted at the Ouelessebougou Clinical Research Unit of the Malaria Research and Training Centre (MRTC) of the University of Bamako in Mali. Before the commencement of screening, the study team met with community leaders, village health workers, and heads of households from each village to explain the study and obtain approval to conduct the study. Village health workers then used a door-to-door approach to inform households of the date and location where consenting and screening would take place. Participants were included in the trial if they met the following criteria: positive for *P. falciparum* gametocytes by microscopy (i.e. ≥1 gametocytes recorded in a thick film against 500 white blood cells, equating to 16 gametocytes/µL with a standard conversion of 8000 white blood cells (WBC)/µL blood); absence of other non-*P. falciparum* species on blood film; haemoglobin density of ≥10 g/dL; G6PD normal (male >4 IU/g Hb, female >6 IU/g Hb); aged between 10-50 years; bodyweight of ≤80 kg; no clinical signs of malaria, defined by fever (≥37·5°C); no signs of acute, severe or chronic disease; consistent with the long half-life of tafenoquine, use of effective contraception for 5 half-lives (3 months) after the end of tafenoquine treatment. Exclusion criteria included pregnancy (tested at enrolment by urine test) or lactation, allergies to any of the study drugs, use of other medication (except for paracetamol and/or aspirin), use of antimalarial drugs over the past week, family history of congenital prolongation of the corrected QT (QTc) interval, current/recent treatment with drugs known to extend QTc interval, and blood transfusion in last 90 days. Prior to screening and prior to study enrolment, participants provided written informed consent (≥18 years) or assent with written parental consent (10-17 years).

Ethical approval was granted by the Ethics Committee of the Faculty of Medicine, Pharmacy, and Dentistry of the University of Science, Techniques, and Technologies of Bamako (Bamako, Mali) (N°2021/189/CE/USTTB), and the Research Ethics Committee of the London School of Hygiene and Tropical Medicine (London, United Kingdom) (LSHTM Ethics Ref: 26257).

### Randomisation and masking

Allocation to four treatment groups (artemether-lumefantrine, artemether-lumefantrine with primaquine [0·25 mg/kg], sulfadoxine-pyrimethamine plus amodiaquine, and sulfadoxine-pyrimethamine plus amodiaquine with tafenoquine [1·66 mg/kg]) was randomised in a 1:1:1:1 ratio. Enrolment continued until 80 participants were enrolled (20 individuals assigned to each treatment group). An independent MRTC statistician randomly generated the treatment assignment using Stata version 16 (StataCorp, College Station, Texas, USA), which was linked to participant ID number. The statistician prepared sealed, opaque envelopes with the participant ID number on the outside and treatment assignment inside which were sent to the MRTC study pharmacist. The study pharmacist provided treatment and was consequently not blinded to treatment assignment; staff involved in assessing safety and laboratory outcomes were blinded.

### Procedures

Artemether-lumefantrine treatment (Coartem, Novartis, Basel, Switzerland) was administered over three days as per manufacturer instructions (appendix pp. 2). A single dose of 0·25 mg/kg primaquine was administered on day 0 in parallel with the first dose of artemether-lumefantrine as described previously (ACE Pharmaceuticals, the Netherlands).^14^ Participants in the sulfadoxine-pyrimethamine plus amodiaquine groups were treated with standard doses of sulfadoxine-pyrimethamine plus amodiaquine (Guilin Pharmaceutical (Shanghai) Co., Ltd. Rm. 701, 780 Cailun Rd., Zhangjiang Hi Tech Park, Shanghai, 201203 China) as per manufacturer instructions (appendix pp. 2). A single dose of tafenoquine (Arakoda, 60° pharmaceuticals LLC, DC, USA) was administered on day 0 in parallel with the first doses of sulfadoxine-pyrimethamine and amodiaquine. Tafenoquine dosing was weight based to standardise efficacy and risk variance (appendix pp. 3). The dose of 1·66 mg/kg chosen for the study was based on good safety profile and efficacy in Malian adults and children ≥12 years old.^17^ G6PD testing was conducted using both semi-quantitative (OSMMR-D G-6-PD test; R&D Diagnostics, Aghia Paraskevi, Greece) and quantitative testing (STANDARD^TM^ G6PD Test, SD BIOSENSOR, Suwon, South Korea); inclusion in the study required normal enzyme function to be determined by both methods.

Participants received a full clinical and parasitological examination on days 2, 5, 7, 14, 21, and 28 after receiving the first dose of the study drugs (appendix pp. 3). Giemsa stained thick film microscopy was performed as described previously, with asexual stages counted against 200 WBC and gametocytes counted against 500 WBC.^14^ For molecular gametocyte quantification, total nucleic acids were extracted using a MagNAPure LC automated extractor (Total Nucleic Acid Isolation Kit-High Performance; Roche Applied Science, Indianapolis, IN, USA). Male and female gametocytes were quantified in a multiplex reverse transcriptase quantitative PCR (RT-qPCR) assay (appendix pp. 4).^21^ Samples were classified as negative for a particular gametocyte sex if the qRT-PCR quantified density of gametocytes of that sex was less than 0·01 gametocytes per μL (i.e. one gametocyte per 100 μL of blood sample). Haemoglobin density was measured using a haemoglobin analyser (HemoCue; AB Leo Diagnostics, Helsingborg, Sweden) or automatic haematology analyser (HumaCount 5D; Wiesbaden, Germany). Additional venous blood samples were taken for biochemical and infectivity assessments on day 0, 2, 5, 7, and 14 in all treatment groups. Aspartate transaminase (AST), alanine transaminase (ALT) and blood creatine levels were determined using automatic biochemistry analyser Human 100 (Wiesbaden, Germany). For each assessment of infectivity, ~75 locally reared female *Anopheles gambiae* mosquitoes were allowed to feed for 15-20 minutes on venous blood samples (Lithium Heparin VACUETTE tube, Greiner Bio-One, Kremsmünster, Austria) through a prewarmed glass membrane feeder system (Coelen Glastechniek, Weldaad, the Netherlands). Surviving mosquitoes were dissected on the 7^th^ day after feeding; midguts were stained in 1% mercurochrome and examined for the presence and density of oocysts by expert microscopists. Adverse events (AE) were graded by the study clinician for severity (mild, moderate, severe) and relatedness to study medication (unrelated or unlikely, possibly, probably, or definitely related). A drop in haemoglobin concentration of ≥40% from baseline was categorised as a haematological severe AE. An external data safety and monitoring committee was assembled prior to the trial, and safety data was discussed after enrolment of 40 participants, and after the final follow-up visit of the last participant.

### Data analysis

Sample size estimation was based on infectivity for participants in previous trials in the same study setting using a mixed effects logistic regression model that accounted for correlation between mosquito observations from the same participant.^13,14,17,18^ With an expected reduction in infectivity of 90% as previously detected for efficacious doses of primaquine and tafenoquine,^14,17,18^ we calculated 92% empirical power to detect >85% reduction in infectivity with a one-tailed test with an α=0·05 level of significance when including 20 participants and dissecting 50 mosquitoes at each time point. When using an α=0·025, empirical power was calculated at 90%.

Clinical and entomological data were double entered into a MS Access database and analysed using STATA (version 16.0) and SAS (version 9.4). Statistical analyses of mosquito infection rate and oocyst density were analysed at timepoints after baseline only for those individuals who were infectious at baseline. The prevalence of gametocytes and infectious individuals were compared within and between treatment groups using one-sided Fishers exact tests. Haemoglobin levels were compared using paired *t* tests (*t* score) for within-group analyses and linear regression adjusted for baseline levels of each measure for between-group analyses (*t* score, coefficient with 95% CI). Percentage change from baseline was analysed using two-way *t* tests. The proportion of gametocytes that were male was analysed for all values with total gametocyte densities of 0·2 gametocytes per μL or more.^14^ Gametocyte circulation time was calculated to determine the mean number of days that a mature gametocyte circulates in the blood before clearance, using a deterministic compartmental model that assumes a constant rate of clearance and has a random effect to account for repeated measures on individuals, as described previously;^22^ difference in circulation time between groups and between gametocyte sexes was analysed using *t* tests (*t* score). Area under the curve (AUC) of gametocyte density per participant over time was calculated using the linear trapezoid method and was analysed by fitting linear regression models to the log_10_ adjusted AUC values, with adjustment for baseline gametocyte density (*t* score, coefficient with 95% CI). All other analyses of quantitative data were done using Wilcoxon sign rank tests (z-score) and Wilcoxon rank-sum tests (z-score). All comparisons were defined before study completion and analyses were not adjusted for multiple comparisons. For all analyses, the threshold for statistical significance was set at p<0·05. The trial is registered with ClinicalTrials.gov, NCT05081089.

## Results

Between 13 Oct and 16 Dec 2021, 1290 individuals were assessed for eligibility and 80 were enrolled and randomly assigned to one of the four treatment groups (n=20 per group) of which 77 (96%) completed all follow-up visits (figure 1). Participant baseline characteristics were similar between the study groups (table 1).

**Figure 1.**
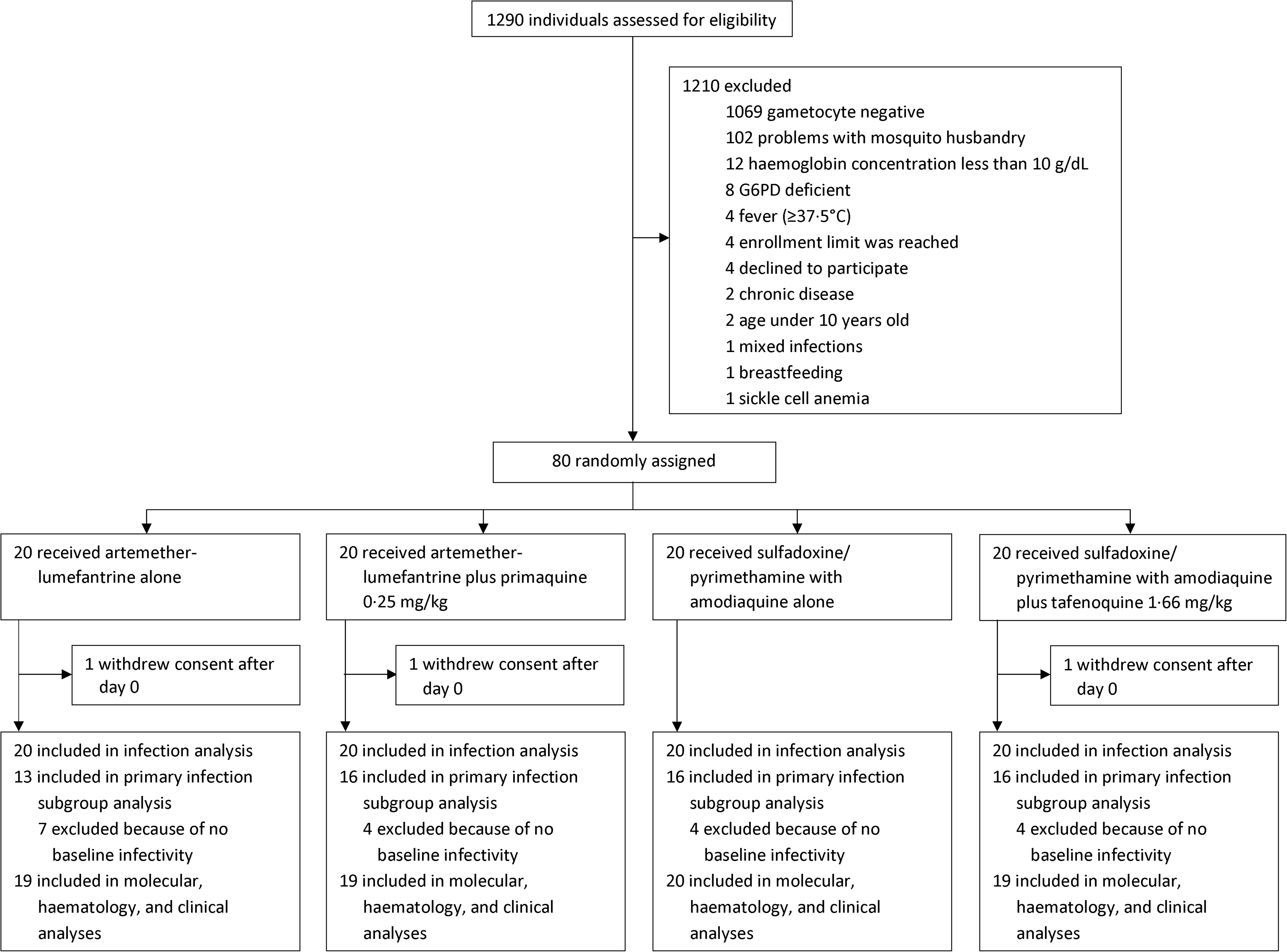
Screening and recruitment profile. 77 of 80 participants completed all follow-up visits: one in the artemether-lumefantrine group, one in the artemether-lumefantrine with primaquine group and one in the sulfadoxine-pyrimethamine plus amodiaquine with tafenoquine group did not complete all visits.

**Table 1.**
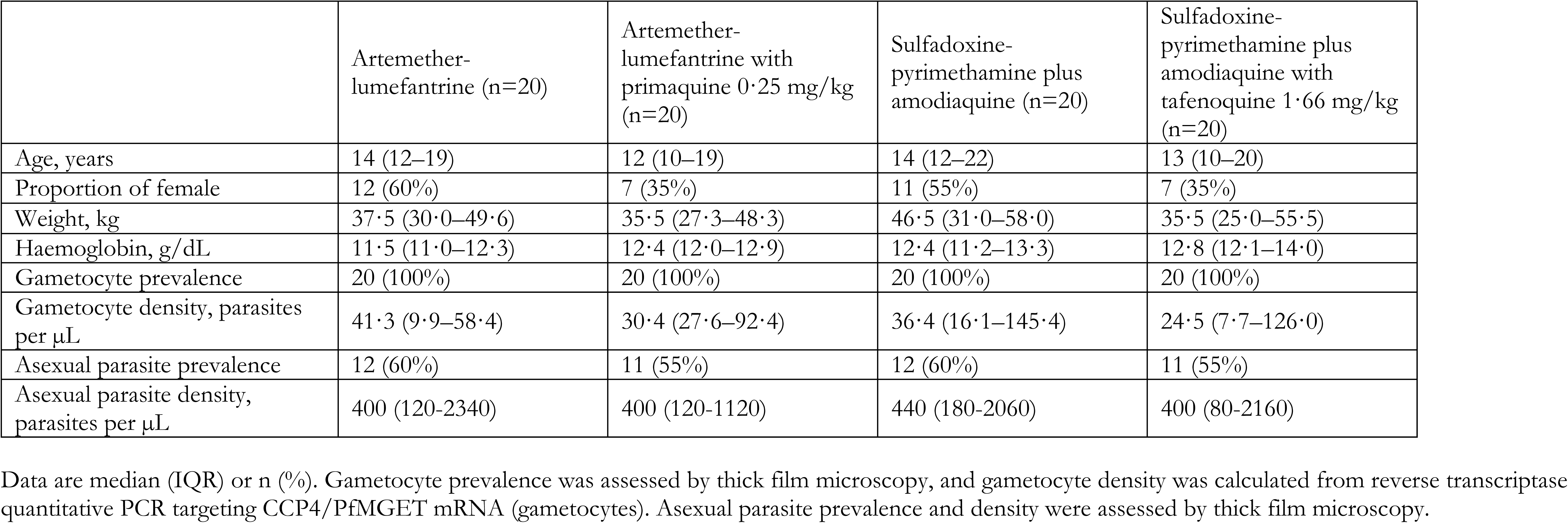
Baseline characteristics.

|  | Artemether-lumefantrine (n=20) | Artemether-lumefantrine with<br>primaquine 0·25 mg/kg<br>(n=20) | Sulfadoxine-pyrimethamine plus<br>amodiaquine (n=20) | Sulfadoxine-pyrimethamine plus<br>amodiaquine with<br>tafenoquine 1·66 mg/kg<br>(n=20) |
| --- | --- | --- | --- | --- |
| Age, years | 14 (12–19) | 12 (10–19) | 14 (12–22) | 13 (10–20) |
| Proportion of female | 12 (60%) | 7 (35%) | 11 (55%) | 7 (35%) |
| Weight, kg | 37·5 (30·0–49·6) | 35·5 (27·3–48·3) | 46·5 (31·0–58·0) | 35·5 (25·0–55·5) |
| Haemoglobin, g/dL | 11·5 (11·0–12·3) | 12·4 (12·0–12·9) | 12·4 (11·2–13·3) | 12·8 (12·1–14·0) |
| Gametocyte prevalence | 20 (100%) | 20 (100%) | 20 (100%) | 20 (100%) |
| Gametocyte density, parasites<br>per µL | 41·3 (9·9–58·4) | 30·4 (27·6–92·4) | 36·4 (16·1–145·4) | 24·5 (7·7–126·0) |
| Asexual parasite prevalence | 12 (60%) | 11 (55%) | 12 (60%) | 11 (55%) |
| Asexual parasite density,<br>parasites per µL | 400 (120–2340) | 400 (120–1120) | 440 (180–2060) | 400 (80–2160) |

Data are median (IQR) or n (%). Gametocyte prevalence was assessed by thick film microscopy, and gametocyte density was calculated from reverse transcriptase quantitative PCR targeting CCP4/PfMGET mRNA (gametocytes). Asexual parasite prevalence and density were assessed by thick film microscopy.

Before treatment, 61 (76%) of 80 participants were infectious to mosquitoes, with a median of 5·5% (IQR 1·4–15·5) of mosquitoes becoming infected (table 2; appendix pp. 5). At day 2, 2 (11%) of 19 participants in the artemether-lumefantrine group and 0 (0%) of 19 participants in the artemether-lumefantrine with primaquine group infected mosquitoes (figure 2). In individuals who were infectious before treatment, the median percentage reduction in mosquito infection rate 2 days after treatment was 100% (IQR 97·2 to 100) for individuals treated with artemether-lumefantrine (n=19; p=0·026) and 100% (IQR 100 to 100) with artemether-lumefantrine with primaquine (n=19; p=0·0001). One participant who received artemether-lumefantrine was infectious on day 5, but not infectious on day 0 or 2. Furthermore, there was one participant in the artemether-lumefantrine with primaquine group who was infectious at baseline and at day 5, but not at day 2. At day 7, 11 (55%) of 20 participants in the sulfadoxine-pyrimethamine plus amodiaquine group and 0 (0%) of 19 participants in the sulfadoxine-pyrimethamine plus amodiaquine with tafenoquine group infected any number of mosquitoes. In individuals who were infectious before treatment, the median percentage reduction in mosquito infection rate 7 days after treatment was 63·6% (IQR 0·62 to 100) for individuals treated with sulfadoxine-pyrimethamine plus amodiaquine (n=11; p=0·0090) compared with 100% (IQR 100 to 100) with sulfadoxine-pyrimethamine plus amodiaquine with tafenoquine (n=19; p<0·0001).

**Figure 2.**
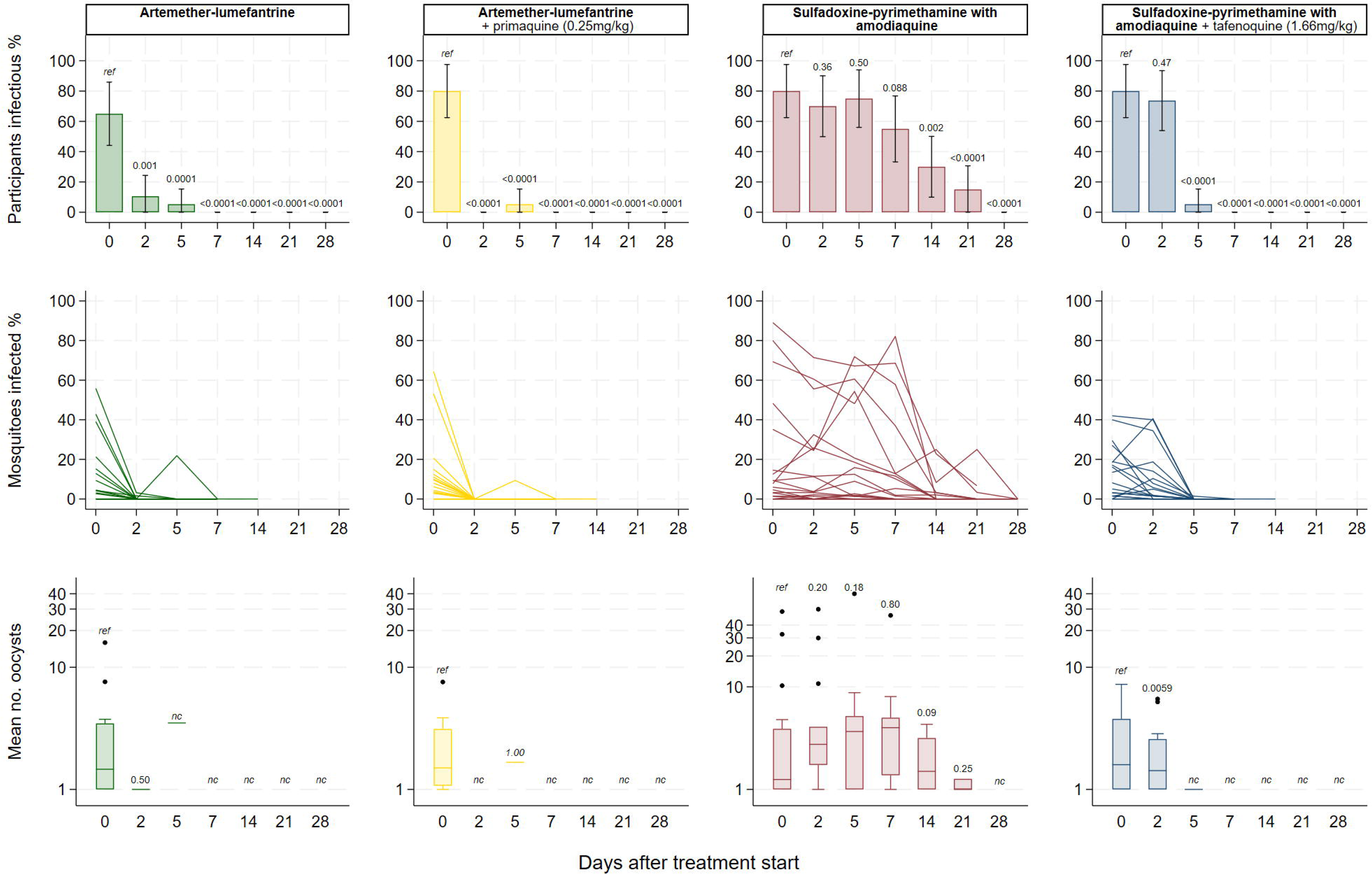
Participant infectivity and proportion of mosquitoes infected in direct membrane feeding assays. Before treatment, 76% of participants were infectious to mosquitoes, infecting a median of 9.7% of mosquitoes. By day 2, the median within-person reduction in mosquito infection rate following AL and AL+PQ was 100%. Transmission-blocking effects in sulfadoxine-pyrimethamine plus amodiaquine showed 19.29% reduction in transmission by day 5 compared to 100% for sulfadoxine-pyrimethamine plus amodiaquine with tafenoquine.

**Table 2.** Infectivity to mosquitoes before and after treatment.

|  | Infectious individuals* | Median mosquito infection rate † (IQR) | Median reduction in mosquito infection rate ‡ (IQR) | P-value § | P-value ¶ |
| --- | --- | --- | --- | --- | --- |
| **Pretreatment** |  |  |  |  |  |
| Artemether-lumefantrine | 13/20 (65%) | 9.38% (4.23 - 21.31) | .. | .. | .. |
| Artemether-lumefantrine with primaquine 0.25 mg/kg | 16/20 (80%) | 9.84% (5.77 - 13.13) | .. | .. | .. |
| Sulfadoxine-pyrimethamine plus amodiaquine | 16/20 (80%) | 9.26% (4.34 - 38.49) | .. | .. | .. |
| Sulfadoxine-pyrimethamine plus amodiaquine with tafenoquine 1.66 mg/kg | 16/20 (80%) | 14.80% (3.20 - 20.97) | .. | .. | .. |
| **Day 2** |  |  |  |  |  |
| Artemether-lumefantrine | 2/19 (11%) | 2.39% (2.00 - 2.78) | 100% (97.2 - 100) | 0.026 | Reference |
| Artemether-lumefantrine with primaquine 0.25 mg/kg | 0/19 (0%) | .. | 100% (100 - 100) | 0.0001 | 0.15 |
| Sulfadoxine-pyrimethamine plus amodiaquine | 14/20 (70%) | 17.98% (3.60 - 30.83) | 30.42% (7.16 - 80.37) | 0.018 | Reference |
| Sulfadoxine-pyrimethamine plus amodiaquine with tafenoquine 1.66 mg/kg | 14/19 (74%) | 6.87% (1.66 - 17.57) | 67.48% (19.80 - 94.17) | 0.055 | 0.33 |
| **Day 5** |  |  |  |  |  |
| Artemether-lumefantrine | 1/19 (5%) | 21.88% (21.88 - 21.88) | 100% (100 - 100) | 0.0005 | Reference |
| Artemether-lumefantrine with primaquine 0.25 mg/kg | 1/19 (5%) | 9.38% (9.38 - 9.38) | 100% (100 - 100) | 0.0065 | 1.0 |
| Sulfadoxine-pyrimethamine plus amodiaquine | 15/20 (75%) | 15.87% (2.26 - 51.22) | 19.29% (48.13 - 70.96) | 0.78 | Reference |
| Sulfadoxine-pyrimethamine plus amodiaquine with tafenoquine 1.66 mg/kg | 1/19 (5%) | 1.45% (1.45 - 1.45) | 100% (100 - 100) | <0.0001 | <0.0001 |
| **Day 7** |  |  |  |  |  |
| Artemether-lumefantrine | 0/19 (0%) | .. | 100% (100 - 100) | 0.0005 | Reference |
| Artemether-lumefantrine with primaquine 0.25 mg/kg | 0/19 (0%) | .. | 100% (100 - 100) | 0.0001 | 1.0 |
| Sulfadoxine-pyrimethamine plus amodiaquine | 11/20 (55%) | 12.77% (7.77 - 47.40) | 63.60% (0.62 - 100) | 0.0090 | Reference |
| Sulfadoxine-pyrimethamine plus amodiaquine with tafenoquine 1.66 mg/kg | 0/19 (0%) | .. | 100% (100 - 100) | <0.0001 | 0.0001 |

\* Individuals were classed as infectious if direct membrane feeding assays resulted in at least one mosquito with any number of oocysts. Mosquito infection measures (percentage infection and oocyst density) are shown for all participants who were infectious at baseline, and oocyst densities are from all infected mosquitoes.
† Median proportion of mosquitoes infected by each participant, where for each participant the mosquito infection rate was the number of mosquitoes infected as a proportion of all mosquitoes surviving to dissection.
‡ Median reduction (relative to baseline) in mosquito infection rate at the given timepoints. All values are for individuals who were infectious to mosquitoes before treatment (ie, infected any number of mosquitoes).
§ Within-group comparison in median reduction in mosquito infection rate (primary outcome).
¶ Between-group comparison in median reduction in mosquito infection rate. Full details of mosquito feeding assay outcomes are in appendix (pp. 5).

The median reduction in mosquito infection rate in the artemether-lumefantrine group was not significantly different from the artemether-lumefantrine with primaquine group at any timepoint. No mosquito infections were observed after day 5 in either artemether-lumefantrine treatment groups. In the sulfadoxine-pyrimethamine plus amodiaquine and sulfadoxine-pyrimethamine plus amodiaquine with tafenoquine groups respectively, 14 (70%) of 20 and 14 (74%) of 19 individuals were infectious to mosquitoes at day 2, while 15 (75%) of 20 and 1 (5%) of 19 were infectious at day 5. Median reduction in mosquito infection rate in individuals infectious at baseline was significantly different between arms by day 5, when there was a 19·29% (IQR 48·13 to 78·96) reduction in the sulfadoxine-pyrimethamine plus amodiaquine group, and near total abrogation of infection with the addition of tafenoquine (100%, IQR 100 to 100) (table 2).

Gametocyte densities declined after initiation of treatment in all treatment groups, although the decrease was much less rapid in the sulfadoxine-pyrimethamine plus amodiaquine group (figure 3; appendix pp. 7-8); 20 (100%) of 20 participants treated with sulfadoxine-pyrimethamine plus amodiaquine alone were still gametocyte positive at the final day of observation (day 28), compared with 11 (58%) of 19 after artemether-lumefantrine alone. Total gametocyte circulation time was estimated at 5·3 days (95% CI 4·5-6·0) in the artemether-lumefantrine group and 2·9 days (2·4-3·3) in the artemether-lumefantrine with primaquine group (appendix pp. 6); the same measure was estimated at 9·1 days (7·3-11·0) and 3·3 days (2·9-3·6) in the sulfadoxine-pyrimethamine plus amodiaquine and sulfadoxine-pyrimethamine plus amodiaquine with tafenoquine groups respectively. Gametocyte sex ratios were initially similar in all treatment groups, but the ratio increased towards male in the artemether-lumefantrine group from day 2 after treatment (median proportion of male gametocytes 0·46 [IQR 0·37-0·51] on day 0 and 0·80 [0·60-0·92] (p=0·013) on day 2) (appendix pp. 7-9). In the sulfadoxine-pyrimethamine plus amodiaquine group, sex ratios remained largely unchanged but with an increasing bias toward females over the 28-day trial period.

**Figure 3.**
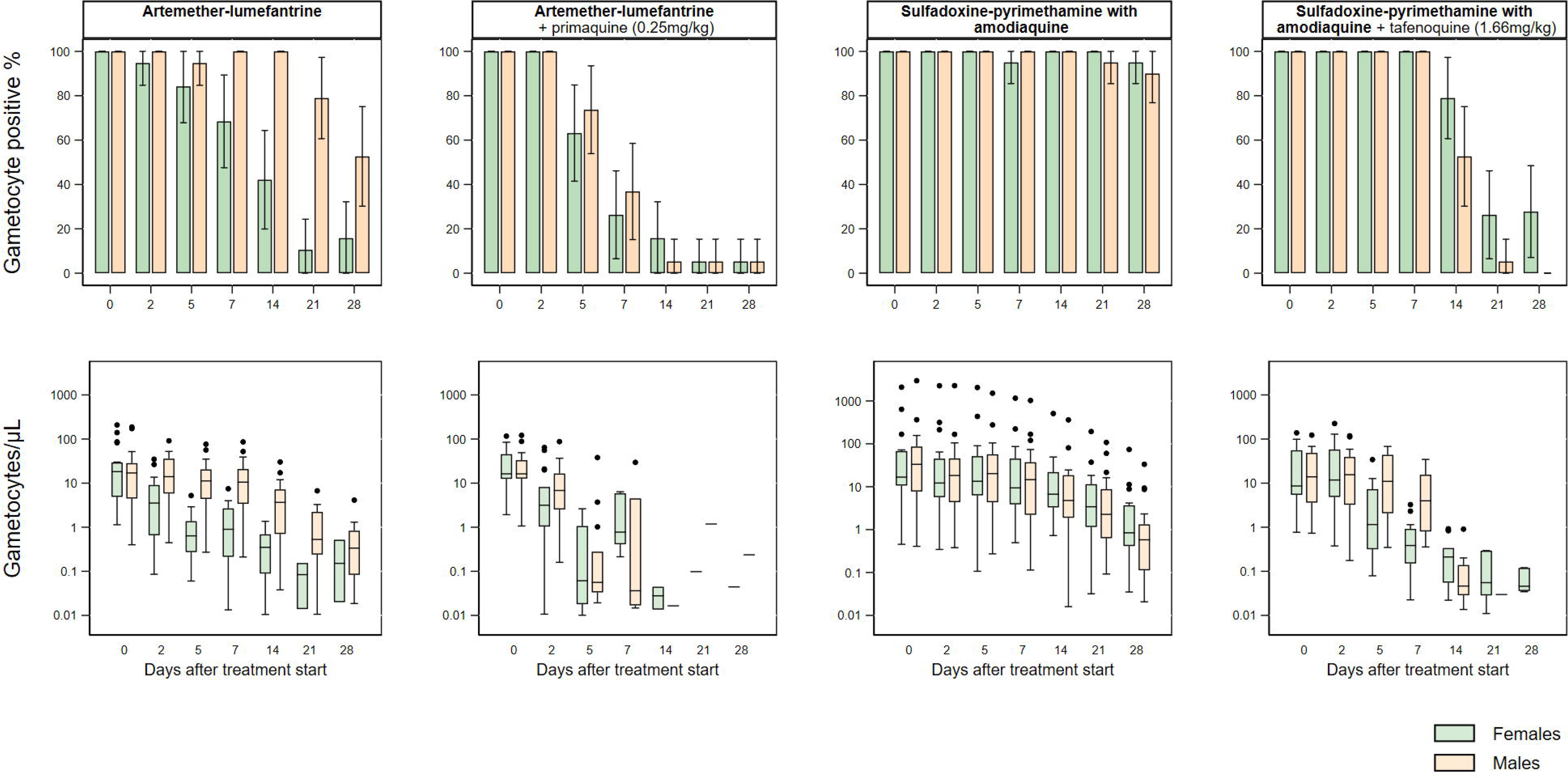
Male and female gametocyte density and prevalence. Gametocyte densities decreased after initiation of treatment in all study groups, although the decrease was less rapid in the sulfadoxine-pyrimethamine plus amodiaquine alone group. Gametocyte sex ratios were initially similar in all treatment groups, but female gametocyte density decreased substantially in the artemether-lumefantrine group by day 5 after treatment, resulting in a significantly male-biased sex ratio.

Treatment with primaquine (p=0·0023) resulted in significantly male-biased ratios from day 2 (p=0·0023), while male bias due to tafenoquine was significant from day 5 (p<0·0001). The absolute infectivity of persisting gametocytes was significantly lower in the sulfadoxine-pyrimethamine plus amodiaquine with tafenoquine group than in the sulfadoxine-pyrimethamine plus amodiaquine group (day 2 odds-ratio (OR) 0·59, p<0·0001; day 5 OR 0·0077, p<0·0001), whereas in the artemether-lumefantrine groups too few gametocytes persisted to allow this comparison (appendix pp. 10).

There were no haemolytic severe adverse events (drop of >40% from baseline). Transient reductions in haemoglobin density were greater in the sulfadoxine-pyrimethamine groups than in the artemether-lumefantrine groups, with significant reductions in haemoglobin density observed at day 1 and 2 (during the period of treatment administration) in both sulfadoxine-pyrimethamine plus amodiaquine groups (mean change between −4·99% and −6·35%); these resolved by day 5 (appendix pp. 11-12). The maximum drop in haemoglobin between baseline and any study timepoint was 15·4% in the artemether-lumefantrine group, 15·9% in the artemether-lumefantrine with primaquine group, 24·3% in the sulfadoxine-pyrimethamine plus amodiaquine group and 23·5% in the sulfadoxine-pyrimethamine plus amodiaquine with tafenoquine group.

Overall, 50 (62·5%) of the 80 participants experienced a total of 92 adverse events during follow-up, of which 61 were at least possibly related to the study drugs. 48 (79%) of the at least possibly related adverse events which were categorised as mild and 13 (21%) as moderate. No grade 3 or serious adverse events occurred (table 3). The most common adverse events were headaches, abdominal pain, and nausea. No severe laboratory abnormalities occurred; all possibly drug related laboratory abnormalities normalised on the subsequent visit (appendix pp. 13).

**Table 3.** Frequency of adverse events.

| Description | All (n=80) | Artemether-lumefantrine (n=20) | Artemether-lumefantrine with | Sulfadoxine-pyrimethamine plus | Sulfadoxine-pyrimethamine plus amodiaquine |
| --- | --- | --- | --- | --- | --- |

|  |  |  | primaquine 0.25 mg/kg (n=20) | amodiaquine (n=20) | with tafenoquine 1.66 mg/kg (n=20) |
| --- | --- | --- | --- | --- | --- |
| **All** | 62.5% (50/80) | 30% (6/20) | 65% (13/20) | 80% (16/20) | 75% (15/20) |
| *P-value* | 0.16* | .. | 0.056** | .. | 1.00** |
| **Mild related AE** | 36% (29/80) | 20% (4/20) | 45% (9/20) | 50% (10/20) | 30% (6/20) |
| *P-value* | 0.37* | .. | 0.176** | .. | 0.333** |
| **Moderate related AE** | 11% (9/80) | 0% (0/20) | 0% (0/20) | 15% (3/20) | 30% (6/20) |
| *P-value* | 0.012* | .. | *nc* | .. | 0.451** |
| **Severe related AE** | 0% (0/80) | 0% (0/20) | 0% (0/20) | 0% (0/20) | 0% (0/20) |
| *P-value* | *nc* | .. | *nc* | .. | *nc* |

Number of participants per group who experienced adverse events. If there were multiple episodes per participant, the highest grade is presented in this table. Classification as ‘related to treatment’ was defined as probably, possibly or definitely related to treatment, as described in the methods. P-values are from Fisher’s exact tests for differences in proportion of individuals with an AE between all groups\* or between AL+PQ group and the (AL) reference group and SPAQ+TQ group and the (SPAQ) reference group\*\*.
*nc* = not calculable.

## Discussion

In this randomised clinical trial, we determined the gametocytocidal and transmission-blocking activities of 1) the most widely used first-line antimalarial, artemether-lumefantrine, with and without the WHO recommended single low-dose of 0·25 mg/kg of primaquine, and 2) the preferred chemo-preventative treatment, sulfadoxine-pyrimethamine plus amodiaquine, with and without a single dose of 1·66 mg/kg tafenoquine. Artemether-lumefantrine alone blocked almost all transmission within 2 days of treatment initiation, whereas sulfadoxine-pyrimethamine plus amodiaquine had limited effect on gametocyte density and prevalence, with significant reductions in infectivity to mosquitoes only observed after 14 days. While the addition of tafenoquine did not prevent transmission at day 2 after treatment initiation, no infected mosquitoes were observed at the next mosquito feeding timepoint on day 5.

Calls for malaria eradication and concerns about the spread of drug resistance have increased interest in the effects of antimalarial drugs on gametocytes and their infectiousness.^23^ Since transmission cannot be reliably predicted from gametocyte densities and direct mosquito transmission outcomes are technically challenging, assessments of post-treatment infectiousness have been few and inconsistent. Previous studies demonstrated that the gametocytocidal effect of artemether-lumefantrine is superior to that of other ACTs like dihydroartemisinin-piperaquine and pyronaridine-artesunate,^24,25^ but were inconclusive on its ability to prevent transmission to mosquitoes early after treatment.^6–10^ In a selected population with high gametocyte densities and confirmed transmission potential prior to treatment, we demonstrate that artemether-lumefantrine exerts a strong gametocytocidal and transmission-blocking effect. Our finding of a median within-person reduction in mosquito infection rate of 100% 2 days after artemether-lumefantrine treatment corroborates findings from clinical trials in Kenya and Gambia.^6,10^ Gametocyte densities also declined after artemether-lumefantrine, total gametocyte density dropped by 44% (38·44 [IQR 9·15-56·47] to 21·61 [7·51-44·01] gametocytes/µL) between baseline and day 2. However, participants were still carrying transmissible densities of gametocytes at day 2,^26^ which suggests that the reduction in infectivity was due to sex ratio distortion and/or a sterilising effect.^4^

The potent transmission-blocking effect artemether-lumefantrine contrasts with pyronaridine-artesunate and dihydroartemisinin-piperaquine for which significant reductions in mosquito infection rate were only seen from days 10 and 7-14 days post-treatment respectively, when tested in the same facilities.^17,18^ Also for individuals treated with sulfadoxine-pyrimethamine plus amodiaquine, considerable and prolonged post-treatment transmission was observed; 69% (11/16) of individuals infectious to mosquitoes at baseline were still infectious a week later, and three weeks after treatment 19% (3/16) remained infectious. These data align with prior observations that show no change in transmission potential in the week following sulfadoxine-pyrimethamine plus amodiaquine treatment.^14^ Male and female gametocyte density declined initially and then increased slightly at day 5; these transient increases in gametocyte density following sulfadoxine-pyrimethamine plus amodiaquine have been observed previously.^7^ Here, 50% (n/N = 10/20) of individuals had more gametocytes at day 5 than at day 2 (compared to 5·3-15·8% in other treatment groups) which corresponded to increased infectiousness over the same time frame. Unlike in the artemether-lumefantrine groups, treatment with sulfadoxine-pyrimethamine plus amodiaquine did not cause any distortion in sex ratio, and though gametocyte prevalence remained at ≥90% throughout the study, densities declined in the majority of into low likely untransmissible levels in the majority of participants by 21 days post treatment.^26^

As expected, treatment the addition of a single low-dose of tafenoquine to sulfadoxine-pyrimethamine plus amodiaquine resulted in significant transmission-blocking activity among study participants. These results are consistent with previous studies in this setting, where the same low-dose of tafenoquine (1·66 mg/kg) added to dihydroartemisinin–piperaquine led to complete transmission blockade at 7 days post-treatment.^17^ Recent studies have shown sub-optimal *P. vivax* relapse rates after tafenoquine radical cure when co-administered with dihydroartemisnin-piperaquine.^27^ The reasons for this are unknown, but the combination appears effective for *P. falciparum* transmission blockade, albeit with a delayed effect compared to primaquine.^17^ The current study, observing a similar timing of the transmission blocking effect when combined with a non-ACT, argues against a putative artemisinin specific antagonism/interaction as the cause of the delayed effect.^27^ In a controlled human malaria infection trial, the transmission-blocking efficacy of a single 50 mg dose of tafenoquine showed modest reduction of mosquito infection rate at day 4 of 35% and 81% by day 7.^28^ This dose is approximately half of that given in the current trial (1·66 mg/kg) which resulted in a median reduction in mosquito infection rate of 100% (IQR 100-100), while participant infectivity dropped from 70% (n/N=14/20) to 10% (n/N=2/20).^17^

Although it is likely to have a prolonged transmission-blocking, there remain concerns for the safely of tafenoquine in G6PD deficient individuals. Testing for G6PD deficiency is required before administration of tafenoquine for its current indications in treatment of *P. vivax* (i.e. 200 mg daily for 3 days or single 300 mg dose).^29^ This study enrolled only G6PD normal individuals, to ensure comparability between groups. We observed no haematological, grade 3, or serious adverse events in any treatment arm, and transient reductions in haemoglobin density that were not significantly different between groups with and without 8-aminoquinonlines. Our results reinforce the previous observations that single low-doses of primaquine and tafenoquine are safe and well tolerated but data for tafenoquine in G6PD deficient individuals are needed.

Because the primary outcome of the current trial was *P. falciparum* transmission and consistent with previous studies, high-density gametocyte carriers were recruited. With the assessment of infectivity before and after treatment, our findings are highly informative on the transmission-reducing activity of the tested drugs among highly infectious individuals. However, different study populations are required for assessments of community level benefits of antimalarials combined with 8-aminoquinolines when given according to WHO guidelines (i.e at clinical presentation) or in mass treatment campaigns, including SMC. Moreover, our finding that single low dose primaquine appears to have little added benefit on reducing transmission when given with artemether-lumefantrine, needs to be considered in the light of possible use scenarios. Alternative ACTs with larger post-treatment transmission potential such as dihydroartemisinin-piperaquine are typically used in community treatment campaigns and the relevance of primaquine is likely to be larger. Moreover, gametocytes in *PfKelch13* mutant infections may preferentially survive artemisinin exposure and infect mosquitoes.^30^ The addition of a non-artemisinin gametocytocide (such as PQ or TQ) to standard treatment may thus be a valuable tool to limit the spread of artemisinin partial resistance in Africa; the WHO malaria policy and advisory group suggested the broader adoption of single low dose primaquine in countries where partial resistance has been detected.^12^

In conclusion, our findings show that artemether-lumefantrine has intrinsic ability to prevent nearly all mosquito infections, even without primaquine, with good safety profile and reveal considerable post-treatment transmission after sulfadoxine-pyrimethamine plus amodiaquine. The addition of a transmission-blocking drug may be beneficial in maximising the community benefit of seasonal malaria chemoprevention.

## Contributors

WS, AM, MS, MM, TB, CD, and AD conceived the study and developed the study protocol. WS, AM, MS, KS, YSi, SMN, AS, OMD, MD, SOM, YSa, SK, YD, SFT, AD, TB, CD, and AD implemented the trial. KL performed molecular analyses. WS, MS and AM verified the raw data. MS, WS and JB analysed the data. WS, AM, MS, RtH, JB, TB, CD, and AD wrote the first draft of the manuscript. All authors read and approved the final manuscript.

## Data sharing

Anonymised data reported in the manuscript will be made available to investigators who provide a methodologically sound proposal to the corresponding author. The protocol is available upon request.

## Data Availability

All data produced in the present study are available upon reasonable request to the authors.

https://clinepidb.org/ce/app/workspace/analyses/DS_d00683a135/new/variables/EUPATH_0000096/EUPATH_0000151

## Acknowledgments

Primaquine tablets were kindly donated by ACE Pharmaceuticals, Zeewolde, the Netherlands. Tafenoquine tablets were kindly donated by 60° Pharmaceuticals Ltd, USA. We would like to thank the study participants and the population of Ouelessebougou, Mali for their cooperation. Finally, we would like to thank the local safety monitor, members of the data safety and monitoring board for their assistance.

