## Appendices for "Artemether-lumefantrine with or without single-dose primaquine and sulfadoxine-pyrimethamine plus amodiaquine with or without single-dose tafenoquine to reduce *Plasmodium falciparum* transmission: a phase 2 single-blind randomised clinical trial in Ouelessebougou, Mali"

**Appendix**

### Supplementary information 1. Antimalarial treatment dosing

A. Artemether-lumefantrine (AL)

Participants in the AL or AL-PQ arm were treated with standard doses of AL from day 0-2. AL treatment tablets containing 20/120 mg artemether/lumefantrine or 80/480 mg artemether/lumefantrine (Coartem, Novartis, Basel, Switzerland) were administered according to weight as per manufacturer guidelines shown below:

| **Body weight (kg)** | **20/120 mg artemether/lumefantrine tablet** | | | **80/480 mg artemether/lumefantrine tablet** | | |
| --- | --- | --- | --- | --- | --- | --- |
|  | **Day 0** | **Day 1** | **Day 2** | **Day 0** | **Day 1** | **Day 2** |
| **5 to < 15 kg** | 2x 1 tablet | 2x 1 tablet | 2x 1 tablet | - | - | - |
| **15 to < 25 kg** | 2x 2 tablets | 2x 2 tablets | 2x 2 tablets | - | - | - |
| **25 to < 35 kg** | 2x 3 tablets | 2x 3 tablets | 2x 3 tablets | - | - | - |
| **≥ 35 kg** | - | - | - | 2x 1 tablet | 2x 1 tablet | 2x 1 tablet |

B. Primaquine (PQ)

Participants in the AL-PQ arm were given a single low dose of 0·25 mg/kg primaquine (ACE Pharmaceuticals, Zeewolde, The Netherlands) as is currently recommended by the World Health Organization. The single dose of PQ was given on day 0 in parallel with the first dose of AL, administered in an aqueous solution, according to a standard operating procedure (SOP) provided by Sanofi as previously done at the study site when PQ was combined with DP or SPAQ.^1,2^

C. Sulfadoxine-pyrimethamine plus amodiaquine (SPAQ)

Participants in the SPAQ and SPAQ-TQ arm were treated with standard doses of SPAQ (Guilin Pharmaceutical, Shanghai, China). SP tablets containing 500 mg sulfadoxine and 25 mg pyrimethamine and AQ tablets containing 150 mg amodiaquine were administered according to weight as per manufacturer guidelines shown below:

| **Body weight** | **500/50 mg sulfadoxine/pyrimethamine tablet** | | |
| --- | --- | --- | --- |
|  | **Day 0** | **Day 1** | **Day 2** |
| **11 to 20 kg** | 1x 1 tablet | 1x 1 tablet | 1x 1 tablet |
| **21 to 30 kg** | 1x 1·5 tablets | 1x 1·5 tablets | 1x 1·5 tablets |
| **31 to 45 kg** | 1x 2 tablets | 1x 2 tablets | 1x 2 tablets |
| **> 45 kg** | 1x 3 tablets | 1x 3 tablets | 1x 3 tablets |

| **Body weight** | **150 mg amodiaquine tablet** | | |
| --- | --- | --- | --- |
|  | **Day 0** | **Day 1** | **Day 2** |
| **15 to 18 kg** | 1x 1·5 tablets | 1x 1 tablet | 1x 1 tablet |
| **19 to 24 kg** | 1x 1·5 tablets | 1x 1·5 tablets | 1x 1·5 tablets |
| **25 to 35 kg** | 1x 2·5 tablets | 1x 2·5 tablets | 1x 2 tablets |
| **36 to 50 kg** | 1x 3 tablets | 1x 3 tablets | 1x 3 tablets |
| **> 50 kg** | 1x 4 tablets | 1x 4 tablets | 1x 3 tablets |

D. Tafenoquine (TQ)

Participants in the SPAQ-TQ arm received a single dose of 1.66 mg/kg tafenoquine (60degrees Pharma, Washington, US) on day 0, following the first dose of SPAQ. A dose of 1.66 mg/kg TQ is equivalent to a 100 mg single dose in a 60 kg adult. The maximum dose chosen for the current study was based on the reported safety profile of TQ doses ≤300 mg, which is similar to that of standard PQ dosing (15 mg daily for 14 days) in adult G6PD heterozygous adult individuals. ^3^ 100mg tafenoquine tablets were available for this study, and were prepared into a 1 mg/mL solution in water for weight-based dosing in 5 kg bands as follows:

| **Weight min** | **Weight max** | **TQ 1 mg/mL total (mL)** | **Water (mL)** | **Masking solution (mL)** |
| --- | --- | --- | --- | --- |
| 30 | 35 | 54·0 | 136·1 | 10 |
| 35·01 | 40 | 62·3 | 127·7 | 10 |
| 40·01 | 45 | 70·6 | 119·4 | 10 |
| 45·01 | 50 | 78·9 | 111·1 | 10 |
| 50·01 | 55 | 87·2 | 102·8 | 10 |
| 55·01 | 60 | 95·5 | 94·5 | 10 |
| 60·01 | 65 | 103·8 | 86·2 | 10 |
| 65·01 | 70 | 112·1 | 77·9 | 10 |
| 70·01 | 75 | 120·4 | 69·6 | 10 |
| 75·01 | 80 | 128·7 | 61·3 | 10 |

### Supplementary figure 1. Schematic representation of sample collection and analysis pipeline

All arms

Venous collection (EDTA vacutainer) ▪ qRT PCR gametocyte quantification

Venous collection (Heparin vacutainer) ▪ Mosquito feeding assay

Finger-prick collection ▪ Thick film microscopy ▪ Haemoglobin density (Hemocue)

Mosquito feeding assay conducted IF either of previous TWO assays infected any mosquitoes

Day 0: Enrolment

Day 1

Day 2

Day 5

Day 7

Day 14

Day 21

Day 28

### Supplementary table 1. Primer sequences and qPCR conditions for PfMGET CCp4 assay

**PfMGET Primer/Probe Sequences**

| **Primers** | **Sequence** |
| --- | --- |
| Primer-FW (5’-3’) | CGGTCCAAATATAAAAATCCTG |
| Primer-RV (5’-3’) | TGTG TAACG TATG ATTCATTTTC |
| Probe (5’-3’) | FAM-CAGCTCCAG CATTAAAACAC-BHQ1 |

**CCp4** **Primer/Probe Sequences**

| **Primers** | **Sequence** |
| --- | --- |
| Primer-FW (5’-3’) | CACATGAATATGAGAATAAAATTG |
| Primer-RV (5’-3’) | TAGGCGAACATGTGGAAAG |
| Probe (5’-3’) | TexasRed-AGCAACAACGGTATGTGCCTTAAAACG-BHQ2 |

Male and female gametocyte quantification was performed as described previously, using a multiplex RT-qPCR assay.^4^ Assays were run using commercial RT-qPCR mixes (Luna® Universal Probe One-Step RT-qPCR Kit, New England Biolabs, Ipswich, MA, USA). FW = Forward primer. RV = Reverse primer.

### Supplementary table 2. Infectivity to mosquitoes

| **Day of follow-up** | **Treatment arm** | **Infectious individuals* n/N (%)** | **P-value**[^§^](https://www.thelancet.com/journals/lanmic/article/PIIS2666-5247(21)00356-6/fulltext) | **P-value**[^¶^](https://www.thelancet.com/journals/lanmic/article/PIIS2666-5247(21)00356-6/fulltext) | **Mosquito infection rate****  **Median % (IQR)** | **P-value**[^§^](https://www.thelancet.com/journals/lanmic/article/PIIS2666-5247(21)00356-6/fulltext) | **P-value**[^¶^](https://www.thelancet.com/journals/lanmic/article/PIIS2666-5247(21)00356-6/fulltext) | **Oocyst density*** Median (IQR)** | **P-value**[^§^](https://www.thelancet.com/journals/lanmic/article/PIIS2666-5247(21)00356-6/fulltext) | **P-value**[^¶^](https://www.thelancet.com/journals/lanmic/article/PIIS2666-5247(21)00356-6/fulltext) |
| --- | --- | --- | --- | --- | --- | --- | --- | --- | --- | --- |
| Day 0 | Overall | 61/80 (76%) | .. | .. | 9·68 (4·23 to 20·69) | .. | .. | 1·50 (1·00–3·46) | .. | .. |
|  | AL | 13/20 (65%) | *Reference* | *Reference* | 9·38% (4·23 to 21·31) | *Reference* | *Reference* | 1·46 (1·00–3·47) | *Reference* | *Reference* |
|  | AL+PQ (0·25 mg/kg) | 16/20 (80%) | *Reference* | 0·24 | 9·84% (5·30 to 13·75) | *Reference* | 0·88 | 1·50 (1·07–3·13) | *Reference* | 0·93 |
|  | SPAQ | 16/20 (80%) | *Reference* | *Reference* | 9·33% (3·98 to 41·79) | *Reference* | *Reference* | 1·25 (1·00–3·89) | *Reference* | *Reference* |
|  | SPAQ+TQ (1·66 mg/kg) | 16/20 (80%) | *Reference* | 0·65 | 14·80% (3·18 to 23·02) | *Reference* | 0·84 | 1·59 (1·00–3·78) | *Reference* | 0·55 |
| Day 2 | AL | 2/19 (11%) | 0·001 | *Reference* | 0% (0 to 0) | 0·0022 | *Reference* | 1·00 (1·00–1·00) | 0.50 | *Reference* |
|  | AL+PQ (0·25 mg/kg) | 0/19 (0%) | <0·0001 | 0·24 | 0% (0 to 0) | 0·0007 | 0·11 | *nc* | *nc* | *nc* |
|  | SPAQ | 14/20 (70%) | 0·36 | *Reference* | 11·36% (2·33 to 29·15) | 0·066 | *Reference* | 3·00 (1·73–4·08) | 0·20 | *Reference* |
|  | SPAQ+TQ (1·66 mg/kg) | 14/19 (74%) | 0·47 | 0·54 | 3·44% (1·52 to 16·39) | 0·056 | 0·33 | 1·25 (1·00–2·58) | 0·0059 | 0·061 |
| Day 5 | AL | 1/19 (5%) | 0·0001 | *Reference* | 0% (0 to 0) | 0·0022 | *Reference* | *nc* | *nc* | *Reference* |
|  | AL+PQ (0·25 mg/kg) | 1/19 (5%) | <0·0001 | 0·76 | 0% (0 to 0) | 0·0015 | 0·37 | 1·67 (1·67–1·67) | 1·00 | *nc* |
|  | SPAQ | 15/20 (75%) | 0·50 | *Reference* | 14·19% (1·41 to 51·22) | 0·72 | *Reference* | 4·00 (1·77–5·19) | 0·18 | *Reference* |
|  | SPAQ+TQ (1·66 mg/kg) | 1/19 (5%) | <0·0001 | <0·0001 | 0% (0 to 0) | 0·0004 | <0·0001 | *nc* | *nc* | *nc* |
| Day 7 | AL | 0/19 (0%) | <0·0001 | *Reference* | 0% (0 to 0) | 0·0022 | *Reference* | *nc* | *nc* | *Reference* |
|  | AL+PQ (0·25 mg/kg) | 0/19 (0%) | <0·0001 | *nc* | 0% (0 to 0) | 0·0007 | *nc* | *nc* | *nc* | *nc* |
|  | SPAQ | 11/20 (55%) | 0·088 | *Reference* | 7·77% (0 to 24·92) | 0·13 | *Reference* | 4·00 (1·38–5·00) | 0·80 | *Reference* |
|  | SPAQ+TQ (1·66mg/kg) | 0/19 (0%) | <0·0001 | 0·0001 | 0% (0 to 0) | 0·0004 | 0·0001 | *nc* | *nc* | *nc* |
| Day 14 | AL | 0/19 (0%) | <0·0001 | *Reference* | .. | *nc* | *Reference* | *nc* | *nc* | *Reference* |
|  | AL+PQ (0·25 mg/kg) | 0/19 (0%) | <0·0001 | *nc* | 0% (0 to 0) | 0·32 | *nc* | *nc* | *nc* | *nc* |
|  | SPAQ | 6/20 (30%) | 0·002 | *Reference* | 0% (0 to 3·23) | 0·0015 | *Reference* | 1·50 (1·00-3·17) | 0·09 | *Reference* |
|  | SPAQ+TQ (1·66 mg/kg) | 0/19 (0%) | <0·0001 | 0·012 | .. | *nc* | *nc* | *nc* | *nc* | *nc* |
| Day 21 | AL | 0/19 (0%) | <0·0001 | *Reference* | .. | *nc* | *Reference* | *nc* | *nc* | *Reference* |
|  | AL+PQ (0·25 mg/kg) | 0/19 (0%) | <0·0001 | *nc* | .. | *nc* | *nc* | *nc* | *nc* | *nc* |
|  | SPAQ | 3/20 (15%) | <0·0001 | *Reference* | 0% (0 to 3·33) | 0·0033 | *Reference* | 1·00 (1·00-1·27) | 0·25 | *Reference* |
|  | SPAQ+TQ (1·66 mg/kg) | 0/19 (0%) | <0·0001 | 0·125 | .. | *nc* | *nc* | *nc* | *nc* | *nc* |
| Day 28 | AL | 0/19 (0%) | <0·0001 | *Reference* | .. | *nc* | *Reference* | *nc* | *nc* | *Reference* |
|  | AL+PQ (0·25 mg/kg) | 0/19 (0%) | <0·0001 | *nc* | .. | *nc* | *nc* | *nc* | *nc* | *nc* |
|  | SPAQ | 0/20 (0%) | <0·0001 | *Reference* | 0% (0 to 0) | 0·043 | *Reference* | *nc* | *nc* | *Reference* |
|  | SPAQ+TQ (1·66mg/kg) | 0/19 (0%) | <0·0001 | nc | .. | *nc* | *nc* | *nc* | *nc* | *nc* |

*Individuals were classed as infectious if direct membrane feeding assays resulted in at least one mosquito with any number of oocysts. Mosquito infection measures (percent infection and oocyst density) are presented for all participants who were infectious at baseline, and oocyst densities are from all infected mosquitoes. **Mosquito infection rate = median percentage of mosquitoes infected by each participant, where for each participant mosquito infection rate the number of mosquitoes infected as a percentage of all mosquitoes surviving to dissection. ***The average oocyst density for each participant was calculated as the mean number of oocysts in infected mosquitoes (i.e., with at least one oocyst). The value presented in the table is the median of all individuals’ average oocyst intensities (a composite figure of all oocysts/all infected mosquitoes is not statistically valid). *nc* = not calculable, no positive observations. .. = not tested. P-value[^§^](https://www.thelancet.com/journals/lanmic/article/PIIS2666-5247(21)00356-6/fulltext) = within group comparison. P-value[^¶^](https://www.thelancet.com/journals/lanmic/article/PIIS2666-5247(21)00356-6/fulltext) = between group comparison (AL+PQ with AL reference group and SPAQ+TQ with SPAQ reference group). AL = artemether-lumefantrine; AL+PQ = artemether-lumefantrine with primaquine; SPAQ = sulfadoxine-pyrimethamine plus amodiaquine; SPAQ+TQ = sulfadoxine-pyrimethamine plus amodiaquine with tafenoquine.

### Supplementary table 3. Gametocyte circulation time and area under the curve

|  | **Treatment group** | **Total gametocytes (CCP4 & PfMGET)** | **P-value*** | **Female gametocytes (CCP4)** | **P-value*** | **Male gametocytes (PfMGET)** | **P-value*** | **P-value^♂♀^** |
| --- | --- | --- | --- | --- | --- | --- | --- | --- |
| **Circulation time**  **Days (95% CI)** | AL | 5·3 (4·5-6·0) | *Reference* | 3·8 (3·1-4·5) | *Reference* | 6·1 (5·1-7·1) | *Reference* | <0·0001 |
|  | AL+PQ (0·25 mg/kg) | 2·9 (2·4-3·3) | <0·0001 | 2·7 (2·2-3·3) | 0·017 | 3·1 (2·4-3·8) | <0·0001 | 0·0014 |
|  | SPAQ | 9·1 (7·3-11·0) | *Reference* | 11·0 (8·1-13·9) | *Reference* | 7·7 (6·4-9·0) | *Reference* | 0·0087 |
|  | SPAQ+TQ (1·66mg/kg) | 3·3 (2·9-3·6) | <0·0001 | 3·7 (3·2-4·2) | <0·0001 | 2·7 (2·3-3·2) | <0·0001 | 0·0004 |
| **AUC; Median (IQR) gametocytes per µL/day** | AL | 11·6 (3·8–17·5) | *Reference* | 1·2 (0·5–3·9) | *Reference* | 9·3 (3·0–16·4) | *Reference* | <0·0001 |
|  | AL+PQ (0·25 mg/kg) | 4·2 (3·2–8·9) | 0·0012 | 1·0 (0·6–3·4) | 0·46 | 2·6 (1·9–5·2) | 0·0005 | <0·0001 |
|  | SPAQ | 17·8 (6·6–61·2) | *Reference* | 8·7 (3·8–29·9) | *Reference* | 10·7 (2·3–29·7) | *Reference* | 0·86 |
|  | SPAQ+TQ (1·66mg/kg) | 6·5 (2·0–27·0) | 0·0091 | 2·1 (0·9–8·3) | <0·0001 | 4·8 (1·0–14·1) | 0·41 | <0·0001 |

Gametocyte circulation time was calculated using a deterministic compartmental model,^5^ and is presented as the model estimate (mean days) with 95% CI. Area under the curve (AUC) of gametocyte density per participant over time was calculated using the linear trapezoid method,^6^ and is presented as the median and IQR of individual AUC values by treatment arm. P-values are for differences in the t-statistic between AL+PQ and SPAQ+TQ treatment groups and the AL and SPAQ reference group, respectively (*), and for between sexes within treatment groups (^♂♀^).

AL = artemether-lumefantrine; AL+PQ = artemether-lumefantrine with primaquine; SPAQ = sulfadoxine-pyrimethamine plus amodiaquine; SPAQ+TQ = sulfadoxine-pyrimethamine plus amodiaquine with tafenoquine.

### Supplementary table 4. Total gametocyte density, prevalence and sex ratio

|  |  | **Total gametocytes (CCP4 & PfMGET)** | | | | | |
| --- | --- | --- | --- | --- | --- | --- | --- |
| **Day of follow-up** | **Treatment arm** | **Median gametocytes/µL (IQR)** | **P-value**[^¶^](https://www.thelancet.com/journals/lanmic/article/PIIS2666-5247(21)00356-6/fulltext) | **Prevalence**  **n/N (%)** | **P-value**[^¶^](https://www.thelancet.com/journals/lanmic/article/PIIS2666-5247(21)00356-6/fulltext) | **Proportion male**  **Median (IQR)** | **P-value**[^¶^](https://www.thelancet.com/journals/lanmic/article/PIIS2666-5247(21)00356-6/fulltext) |
| Day 0 | *Overall* | 30·44 (16·26–95·35) | .. | 78/80 (98%) | .. | 0·49 (0·40–0·56) | .. |
|  | AL | 38·44 (9·15-56·47) | *Reference* | 20/20 (100%) | *Reference* | 0·46 (0·37–0·51) | *Reference* |
|  | AL+PQ (0·25 mg/kg) | 31·46 (27·86-77·43) | 0·15 | 20/20 (100%) | *nc* | 0·50 (0·37–0·59) | 0·29 |
|  | SPAQ | 53·03 (16·50-147·06) | *Reference* | 18/20 (90%) | *Reference* | 0·53 (0·45–0·66) | *Reference* |
|  | SPAQ+TQ (1·66 mg/kg) | 23·16 (9·01-110·65) | 0·37 | 20/20 (100%) | 0·244 | 0·48 (0·43–0·54) | 0·20 |
| Day 2 | AL | 21·61 (7·51–44·01) | *Reference* | 19/19 (100%) | *Reference* | 0·80 (0·60–0·92) | *Reference* |
|  | AL+PQ (0·25 mg/kg) | 9·97 (3·24–19·62) | 0·24 | 19/19 (100%) | *nc* | 0·65 (0·47–0·79) | 0·057 |
|  | SPAQ | 30·35 (12·49–81·57) | *Reference* | 20/20 (100%) | *Reference* | 0·49 (0·36–0·58) | *Reference* |
|  | SPAQ+TQ (1·66 mg/kg) | 27·27 (9·61–103·67) | 0·83 | 19/19 (100%) | *nc* | 0·48 (0·34–0·57) | 0·79 |
| Day 5 | AL | 11·15 (4·66–21·14) | *Reference* | 18/19 (95%) | *Reference* | 0·96 (0·87–0·98) | *Reference* |
|  | AL+PQ (0·25 mg/kg) | 0·05 (0·02–0·37) | 0·0001 | 16/19 (84%) | 0·30 | 0·67 (0·27–0·89) | 0·0023 |
|  | SPAQ | 31·20 (11·43–111·59) | *Reference* | 20/20 (100%) | *Reference* | 0·49 (0·38–0·57) | *Reference* |
|  | SPAQ+TQ (1·66 mg/kg) | 12·86 (2·89–56·40) | 0·063 | 19/19 (100%) | *nc* | 0·77 (0·65–0·94) | <0·0001 |
| Day 7 | AL | 12·63 (3·40–20·88) | *Reference* | 19/19 (100%) | *Reference* | 0·98 (0·87–1·00) | *Reference* |
|  | AL+PQ (0·25 mg/kg) | 0 (0–0·21) | 0·025 | 8/19 (42%) | 0·0001 | 0·40 (0·08–0·43) | 0·0003 |
|  | SPAQ | 22·65 (6·81–86·89) | *Reference* | 20/20 (100%) | *Reference* | 0·47 (0·31–0·62) | *Reference* |
|  | SPAQ+TQ (1·66 mg/kg) | 4·76 (1·02–15·78) | 0·010 | 18/19 (95%) | 0·49 | 0·91 (0·85–0·96) | <0·0001 |
| Day 14 | AL | 3·69 (0·70–7·20) | *Reference* | 19/19 (100%) | *Reference* | 1·00 (0·97–1·00) | *Reference* |
|  | AL+PQ (0·25 mg/kg) | 0 (0–0) | 0·030 | 4/19 (21%) | <0·0001 | *nc* | *nc* |
|  | SPAQ | 11·30 (5·75–38·27) | *Reference* | 20/20 (100%) | *Reference* | 0·40 (0·29–0·54) | *Reference* |
|  | SPAQ+TQ (1·66 mg/kg) | 0·08 (0·03–0·39) | 0·0004 | 16/19 (84%) | 0·11 | 0·24 (0·04–0·43) | 0·099 |
| Day 21 | AL | 0·40 (0·01–1·56) | *Reference* | 15/19 (79%) | *Reference* | 1·00 (1·00–1·00) | *Reference* |
|  | AL+PQ (0·25 mg/kg) | 0(0–0) | 0·87 | 1/19 (5%) | <0·0001 | 0·92 (0·92–0·92) | 0·14 |
|  | SPAQ | 4·77 (2·06–17·66) | *Reference* | 20/20 (100%) | *Reference* | 0·40 (0·20–0·57) | *Reference* |
|  | SPAQ+TQ (1·66 mg/kg) | 0 (0–0·03) | 0·004 | 6/19 (32%) | <0·0001 | 0 (0–0) | 0·0095 |
| Day 28 | AL | 0·02 (0–0·72) | *Reference* | 11/19 (58%) | *Reference* | 1·00 (0·79–1·00) | *Reference* |
|  | AL+PQ (0·25 mg/kg) | 0 (0–0) | 0·75 | 2/19 (11%) | 0·003 | 1·00 (1·00–1·00) | 1·0 |
|  | SPAQ | 1·50 (0·50–3·92) | *Reference* | 20/20 (100%) | *Reference* | 0·24 (0·11–0·48) | *Reference* |
|  | SPAQ+TQ (1·66 mg/kg) | 0 (0–0·03) | 0·12 | 5/19 (26%) | <0·0001 | *nc* | *nc* |

P-values are for differences between AL+PQ and the AL reference group and SPAQ+TQ and the SPAQ reference group. Density was compared using regression analyses of log10 transformed density values, with adjustment for baseline densities. Prevalence was compared with one sided Fishers exact tests. For males and females, proportion male is given for participants/time-points with total gametocyte densities of 0·2/µL and over, as described previously (1). For the calculation of gametocyte prevalence, samples were classified as negative for a particular gametocyte sex if the estimated density of in gametocytes of that sex was less than 0·01/μL (i.e. one gametocyte per 100 μL of blood sample). P-value[^§^](https://www.thelancet.com/journals/lanmic/article/PIIS2666-5247(21)00356-6/fulltext) = within group comparison. P-value[^¶^](https://www.thelancet.com/journals/lanmic/article/PIIS2666-5247(21)00356-6/fulltext) = between group comparison (AL+PQ with AL reference group and SPAQ+TQ with SPAQ reference group). *nc* = not calculable, no observations/no observations over the threshold density for analysis. .. = not tested. AL = artemether-lumefantrine; AL+PQ = artemether-lumefantrine with primaquine; SPAQ = sulfadoxine-pyrimethamine plus amodiaquine; SPAQ+TQ = sulfadoxine-pyrimethamine plus amodiaquine with tafenoquine.

### Supplementary table 5. Female (CCP4) and male (PfMGET) gametocyte density and prevalence

|  | | **Female gametocytes (CCP4)** | | | | **Male gametocytes (PfMGET)** | | | |
| --- | --- | --- | --- | --- | --- | --- | --- | --- | --- |
| **Day of follow-up** | **Treatment arm** | **Median/µL (IQR)** | **P-value** | **Prevalence**  **n/N (%)** | **P-value** | **Median/µL (IQR)** | **P-value** | **Prevalence**  **n/N (%)** | **P-value** |
| Day 0 | *Overall* | 15·19 (7·32–45·62) | ·· | 78/80 (98%) | ·· | 15·69 (5·46–45·85) | .. | 78/80 (98%) | .. |
|  | AL | 18·18 (4·89–29·44) | *Reference* | 20/20 (100%) | *Reference* | 16·97 (4·47–28·31) | *Reference* | 20/20 (100%) | *Reference* |
|  | AL+PQ (0·25 mg/kg) | 16·22 (12·52–45·29) | 0·19 | 20/20 (100%) | *nc* | 16·22 (12·66–33·14) | 0·17 | 20/20 (100%) | *nc* |
|  | SPAQ | 16·75 (10·65–67·75) | *Reference* | 18/20 (90%) | *Reference* | 33·27 (7·75–86·13) | *Reference* | 18/20 (90%) | *Reference* |
|  | SPAQ+TQ (1·66 mg/kg) | 8·61 (5·42–55·57) | 0·68 | 20/20 (100%) | *nc* | 13·90 (3·62–48·48) | 0·22 | 20/20 (100%) | *nc* |
| Day 2 | AL | 2·77 (0·55–9·07) | *Reference* | 18/19 (95%) | *Reference* | 14·01 (5·82–35·79) | *Reference* | 19/19 (100%) | *Reference* |
|  | AL+PQ (0·25 mg/kg) | 3·15 (1·03–8·18) | 0·60 | 19/19 (100%) | 0.50 | 6·82 (2·52–16·50) | 0·065 | 19/19 (100%) | *nc* |
|  | SPAQ | 12·16 (5·72–44·93) | *Reference* | 20/20 (100%) | *Reference* | 18·26 (4·37–45·51) | *Reference* | 20/20 (100%) | *Reference* |
|  | SPAQ+TQ (1·66 mg/kg) | 11·75 (4·85–57·78) | 0·65 | 19/19 (100%) | *nc* | 15·52 (3·24–39·11) | 0·79 | 19/19 (100%) | *nc* |
| Day 5 | AL | 0·45 (0·08–1·27) | *Reference* | 16/19 (84%) | *Reference* | 9·68 (3·67–20·43) | *Reference* | 18/19 (95%) | *Reference* |
|  | AL+PQ (0·25 mg/kg) | 0·02 (0–0·24) | 0·60 | 12/19 (63%) | 0.14 | 0·04 (0–0·28) | <0·0001 | 14/19 (74%) | 0·090 |
|  | SPAQ | 13·40 (6·29–51·42) | *Reference* | 20/20 (100%) | *Reference* | 20·21 (4·34–56·84) | *Reference* | 20/20 (100%) | *Reference* |
|  | SPAQ+TQ (1·66 mg/kg) | 1·15 (0·32–7·29) | 0·0001 | 19/19 (100%) | *nc* | 10·97 (2·08–43·71) | 0·39 | 19/19 (100%) | *nc* |
| Day 7 | AL | 0·21 (0–1·04) | *Reference* | 13/19 (68%) | *Reference* | 10·51 (3·39–20·88) | *Reference* | 19/19 (100%) | *Reference* |
|  | AL+PQ (0·25 mg/kg) | 0 (0–0·21) | 0·44 | 5/19 (26%) | 0.011 | 0 (0–0·03) | 0·027 | 7/19 (37%) | <0·0001 |
|  | SPAQ | 9·09 (3·78–37·22) | *Reference* | 19/20 (95%) | *Reference* | 14·62 (2·21–37·16) | *Reference* | 20/20 (100%) | *Reference* |
|  | SPAQ+TQ (1·66 mg/kg) | 0·38 (0·15–0·91) | <0·0001 | 18/19 (95%) | 0.53 | 3·97 (0·80–15·47) | 0·14 | 18/19 (95%) | *nc* |
| Day 14 | AL | 0 (0–0·32) | *Reference* | 8/19 (42%) | *Reference* | 3·65 (0·70–7·20) | *Reference* | 19/19 (100%) | *Reference* |
|  | AL+PQ (0·25 mg/kg) | 0 (0–0) | 0·60 | 3/19 (16%) | 0.076 | 0 (0–0) | 0·31 | 1/19 (0·1%) | <0·0001 |
|  | SPAQ | 6·68 (3·30–21·88) | *Reference* | 20/20 (100%) | *Reference* | 4·78 (1·89–18·54) | *Reference* | 20/20 (100%) | *Reference* |
|  | SPAQ+TQ (1·66 mg/kg) | 0·08 (0·02–0·26) | <0·0001 | 15/19 (79%) | 0.047 | 0·01 (0–0·06) | 0·0003 | 10/19 (53%) | 0·0004 |
| Day 21 | AL | 0 (0–0) | *Reference* | 2/19 (0·1%) | *Reference* | 0·40 (0·01–1·56) | *Reference* | 15/19 (79%) | *Reference* |
|  | AL+PQ (0·25 mg/kg) | 0 (0–0) | *nc* | 1/19 (0·1%) | 0.50 | 0 (0–0) | 0·77 | 1/19 (0·1%) | <0·0001 |
|  | SPAQ | 3·41 (1·15–11·43) | *Reference* | 20/20 (100%) | *Reference* | 2·18 (0·47–6·89) | *Reference* | 19/20 (95%) | *Reference* |
|  | SPAQ+TQ (1·66 mg/kg) | 0 (0–0·01) | 0·020 | 5/19 (26%) | <0.0001 | 0 (0–0) | 0·38 | 1/19 (0·1%) | <0·0001 |
| Day 28 | AL | 0 (0–0) | *Reference* | 3/19 (16%) | *Reference* | 0·019 (0–0·42) | *Reference* | 10/19 (53%) | *Reference* |
|  | AL+PQ (0·25 mg/kg) | 0 (0–0) | *nc* | 1/19 (0·1%) | 0.30 | 0 (0–0) | 0·78 | 1/19 (0·1%) | 0·002 |
|  | SPAQ | 0·81 (0·35–3·16) | *Reference* | 19/20 (95%) | *Reference* | 0·47 (0·07–1·16) | *Reference* | 18/20 (90%) | *Reference* |
|  | SPAQ+TQ (1·66 mg/kg) | 0 (0–0·03) | 0·20 | 5/19 (26%) | <0.0001 | 0 (0–0) | *nc* | 0/19 (0%) | <0·0001 |

P-values are for differences between AL+PQ and SPAQ+TQ groups and the reference AL and SPAQ group, respectively. Density was compared using regression analyses of log10 transformed density values, with adjustment for baseline densities. Prevalence was compared with one sided Fishers exact tests. For the calculation of gametocyte prevalence, samples were classified as negative for a particular gametocyte sex if the estimated density of in gametocytes of that sex was less than 0·01 gametocytes per μL (i.e. one gametocyte per 100 μL of blood sample). - = not tested. *ref* = reference group.

AL = artemether-lumefantrine; AL+PQ = artemether-lumefantrine with primaquine; SPAQ = sulfadoxine-pyrimethamine plus amodiaquine; SPAQ+TQ = sulfadoxine-pyrimethamine plus amodiaquine with tafenoquine.

### Supplementary figure 2. Proportion of gametocytes that were male

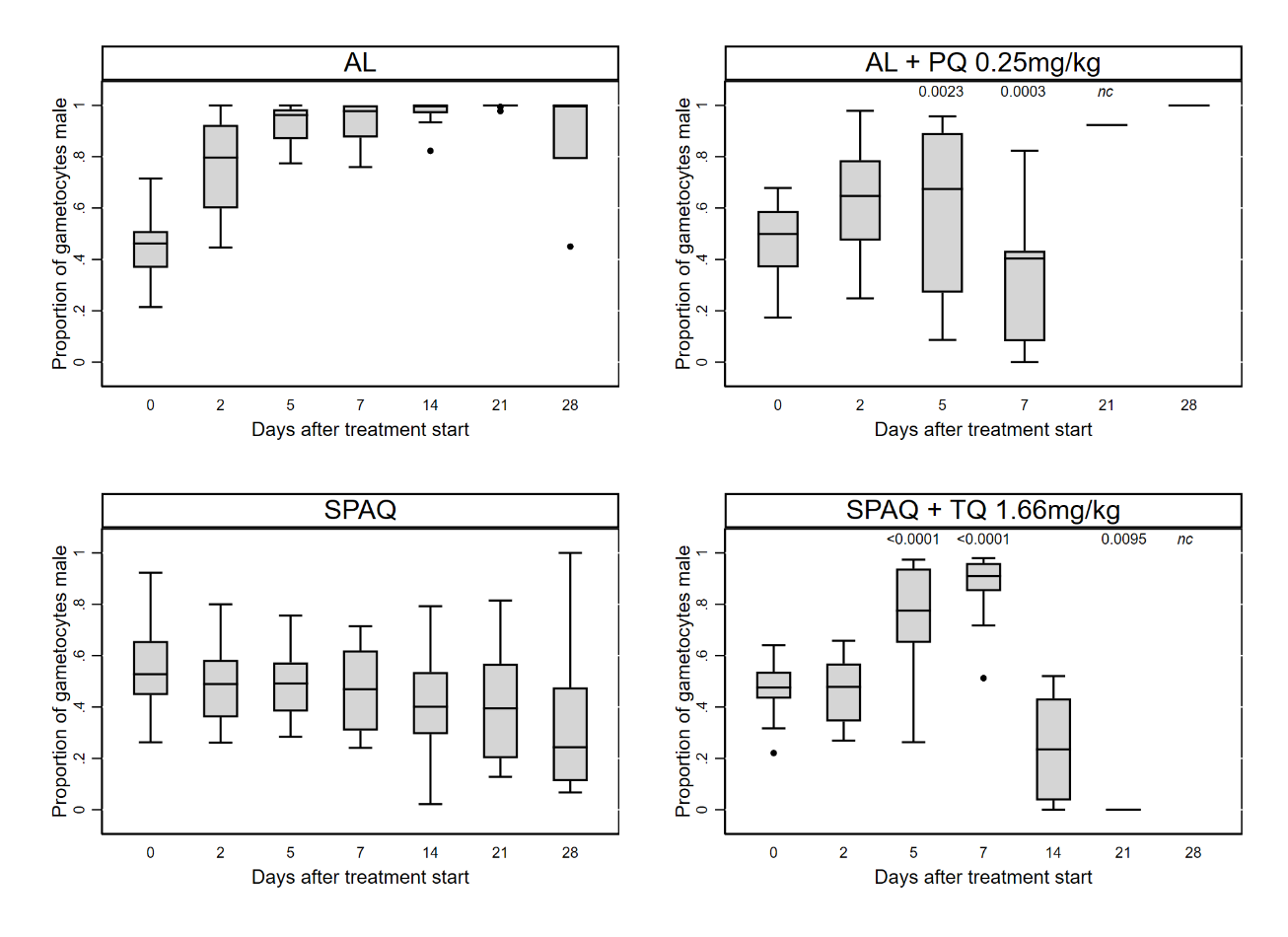

The proportion of gametocytes that were male was calculated for all values with total gametocyte densities of 0·2/µL and over, as described previously.^1^ P-values (<0·05) for differences between treatment groups AL+PQ and SPAQ+TQ and the reference groups AL and SPAQ, respectively, were calculated using Wilcoxon rank sum tests.

**Supplementary table 6. Gametocyte infectivity**

| **Day of follow-up** | **Treatment arm** | **Odds ratio (95% CI)** | **P-value** |
| --- | --- | --- | --- |
| Day 0 | AL | 1 | *Reference* |
|  | AL+PQ (0·25 mg/kg) | 1·16 | *0·289* |
|  | SPAQ | 1 | *Reference* |
|  | SPAQ+TQ (1·66 mg/kg) | 0·95 | *0·733* |
| Day 2 | AL | 1 | *Reference* |
|  | AL+PQ (0·25 mg/kg) | 1·11 | *nc* |
|  | SPAQ | 1 | *Reference* |
|  | SPAQ+TQ (1·66 mg/kg) | 0·59 | <0·0001 |
| Day 5 | AL | 1 | *Reference* |
|  | AL+PQ (0·25 mg/kg) | *nc* | *nc* |
|  | SPAQ | 1 | *Reference* |
|  | SPAQ+TQ (1·66 mg/kg) | 0·0077 | <0·0001 |
| Day 7 | AL | 1 | *Reference* |
|  | AL+PQ (0·25 mg/kg) | 1·0 | *nc* |
|  | SPAQ | 1 | *Reference* |
|  | SPAQ+TQ (1·66 mg/kg) | <0·0001 | *nc* |

Log odds ratios are for the change in mosquito infection rate in the AL+PQ arm compared to the reference (AL) arm and the SPAQ+TQ arm compared to the reference (SPAQ) arm with adjustment for female and male gametocyte densities. *nc* = not calculable, no observations (too few infected mosquitoes for convergence). AL = artemether-lumefantrine; AL+PQ = artemether-lumefantrine with primaquine; SPAQ = sulfadoxine-pyrimethamine plus amodiaquine; SPAQ+TQ = sulfadoxine-pyrimethamine plus amodiaquine with tafenoquine.

### Supplementary table 7. Haemoglobin density and change

Haemoglobin density and percent change in haemoglobin density (relative to baseline) were compared within treatment arms (p-value[^§^](https://www.thelancet.com/journals/lanmic/article/PIIS2666-5247(21)00356-6/fulltext)) using paired t-tests (with day 0 as reference for percent change) and between treatment arms (p-value[^¶^](https://www.thelancet.com/journals/lanmic/article/PIIS2666-5247(21)00356-6/fulltext)) using linear regression (for density, adjusted for baseline Hb density) or two-way t-tests (for percent reduction). AL = artemether-lumefantrine; AL+PQ = artemether-lumefantrine with primaquine; SPAQ = sulfadoxine-pyrimethamine plus amodiaquine; SPAQ+TQ = sulfadoxine-pyrimethamine plus amodiaquine with tafenoquine.

| **Day of follow-up** | **Treatment arm** | **Haemoglobin**  **Mean g/dL (range)** | **P-value**[^§^](https://www.thelancet.com/journals/lanmic/article/PIIS2666-5247(21)00356-6/fulltext) | **P-value**[^¶^](https://www.thelancet.com/journals/lanmic/article/PIIS2666-5247(21)00356-6/fulltext) | **Percent change from day 0** | | **P-value**[^§^](https://www.thelancet.com/journals/lanmic/article/PIIS2666-5247(21)00356-6/fulltext) | **P-value**[^¶^](https://www.thelancet.com/journals/lanmic/article/PIIS2666-5247(21)00356-6/fulltext) |
| --- | --- | --- | --- | --- | --- | --- | --- | --- |
|  |  |  |  |  | **Mean (lower/upper 95% CI)** | **Range** |  |  |
| Day 0 | *Overall* | 12·3 (10·1–15·7) | .. | .. | .. | .. | .. | .. |
|  | AL | 11·5 (10·1–13·2) | *Reference* | *Reference* | .. | .. | .. | .. |
|  | AL+PQ (0·25 mg/kg) | 12·7 (10·7–15·7) | *Reference* | 0·26 | .. | .. | .. | .. |
|  | SPAQ | 12·4 (10·2–14·6) | *Reference* | *Reference* | .. | .. | .. | .. |
|  | SPAQ+TQ (1·66 mg/kg) | 12·8 (10·4–15·2) | *Reference* | 0·85 | .. | .. | .. | .. |
| Day 1 | AL | 11·4 (10·0–13·4) | 0·60 | *Reference* | -0·65 (-4·40/3·10) | -13·64/19·8 | 0·75 | *Reference* |
|  | AL+PQ (0·25 mg/kg) | 12·3 (10·4–15·8) | 0·051 | 0·26 | -2·75 (-5·30/-0·21) | -13·04/5·71 | 0·054 | 0·38 |
|  | SPAQ | 11·6 (9·9–14·0) | 0·0012 | *Reference* | -5·97 (-8·34/-3·59) | -16·94/1·55 | 0·0001 | *Reference* |
|  | SPAQ+TQ (1·66 mg/kg) | 11·9 (10·1–13·9) | 0·003 | 0·67 | -6·35 (-9·95/-2·75) | -23·53/6·73 | 0·003 | 0·87 |
| Day 2 | AL | 11·3 (9·9–13·1) | 0·18 | *Reference* | 1·81 (-1·02/4·65) | -15·84/12·88 | 0·24 | *Reference* |
|  | AL+PQ (0·25 mg/kg) | 12·5 (11·2–15·8) | 0·27 | 0·89 | 1·21 (-1·20/3·61) | -14·29/10·40 | 0·35 | 0·76 |
|  | SPAQ | 11·7 (10·1–13·6) | <0·0001 | *Reference* | 5·01 (3·24/6·77) | -3·45/13·49 | <0·0001 | *Reference* |
|  | SPAQ+TQ (1·66 mg/kg) | 12·1 (10·2–14·3) | 0·001 | 0·81 | 4·99 (2·49/7·48) | -4·42/15·50 | 0·001 | 0·99 |
| Day 5 | AL | 11·4 (10·1–12·5) | 0·39 | *Reference* | -1·07 (-4·25/2·11) | -15·15/18·81 | 0·53 | *Reference* |
|  | AL+PQ (0·25 mg/kg) | 12·5 (11·0–14·9) | 0·50 | 0·76 | -0·74 (-3·14/1·66) | -11·20/14·29 | 0·57 | 0·87 |
|  | SPAQ | 12·1 (10·5–14·6) | 0·034 | *Reference* | -1·89 (-3·47/-0·30) | -7·26/4·29 | 0·35 | *Reference* |
|  | SPAQ+TQ (1·66 mg/kg) | 12·4 (10·6–14·2) | 0·033 | 0·53 | -2·95 (-5·61/-0·29) | -12·88/8·65 | 0·048 | 0·51 |
| Day 7 | AL | 11·6 (9·9–12·9) | 0·83 | *Reference* | 0·71 (-2·62/4·05) | -12·88/21·78 | 0·69 | *Reference* |
|  | AL+PQ (0·25 mg/kg) | 12·3 (10·3–14·9) | 0·004 | 0·24 | -3·15 (-4·98/-1·32) | -10·40/5·04 | 0·004 | 0·060 |
|  | SPAQ | 12·3 (10·6–14·6) | 0·81 | *Reference* | -0·18 (-3·88/3·51) | -24·29/11·43 | 0·93 | *Reference* |
|  | SPAQ+TQ (1·66 mg/kg) | 12·6 (10·8–14·7) | 0·25 | 0·23 | -1·71 (-4·87/1·45) | -13·64/15·75 | 0·31 | 0·55 |
| Day 14 | AL | 12·1 (11·1–14·4) | 0·015 | *Reference* | 5·53 (1·77/9·29) | -8·33/25·74 | 0·012 | *Reference* |
|  | AL+PQ (0·25 mg/kg) | 12·6 (11·0–15·0) | 0·89 | 0·76 | -0·11 (-2·34/2·12) | -9·76/5·71 | 0·93 | 0·019 |
|  | SPAQ | 12·6 (11·1–15·0) | 0·12 | *Reference* | 2·26 (-0·32/4·83) | -11·90/16·19 | 0·11 | *Reference* |
|  | SPAQ+TQ (1·66 mg/kg) | 12·5 (11·0–14·8) | 0·037 | 0·52 | -1·96 (-3·83/-0·10) | -8·82/5·77 | 0·060 | 0·017 |
| Day 21 | AL | 12·2 (11·0–13·6) | 0·024 | *Reference* | 5·99 (1·75/10·24) | -15·38/23·76 | 0·015 | *Reference* |
|  | AL+PQ (0·25 mg/kg) | 12·7 (11·1–15·0) | 0·75 | 0·49 | 1·11 (-2·62/4·83) | -15·92/13·39 | 0·58 | 0·11 |
|  | SPAQ | 12·6 (11·1–14·6) | 0·046 | *Reference* | 2·68 (0·31/5·04) | -2·29/20·95 | 0·044 | *Reference* |
|  | SPAQ+TQ (1·66 mg/kg) | 13·1 (11·4–15·8) | 0·18 | 0·63 | 2·40 (-0·57/5·38) | -13·64/13·46 | 0·14 | 0·89 |
| Day 28 | AL | 12·0 (10·8–13·3) | 0·077 | *Reference* | 4·32 (0·25/8·39) | -11·36/25·74 | 0·058 | *Reference* |
|  | AL+PQ (0·25 mg/kg) | 13·1 (11·5–15·4) | 0·008 | 1·00 | 3·9 (1·44/6·37) | -6·40/10·28 | 0·007 | 0·87 |
|  | SPAQ | 12·5 (10·0–14·5) | 0·66 | *Reference* | 1·18 (-2·40/4·77) | -11·50/20·59 | 0·54 | *Reference* |
|  | SPAQ+TQ (1·66 mg/kg) | 12·9 (11·2–15·2) | 0·70 | 0·46 | 0·91 (-2·15/3·97) | -16·18/11·63) | 0·58 | 0·91 |

### Supplementary figure 3. Haemoglobin density

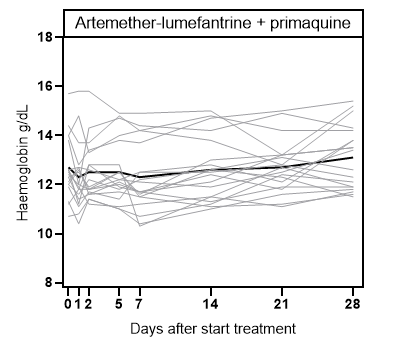

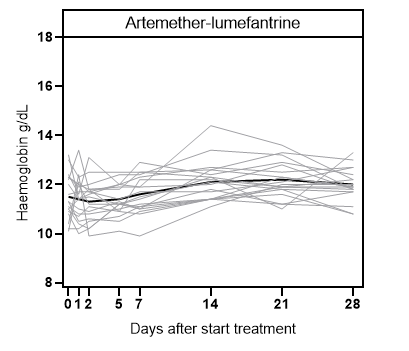

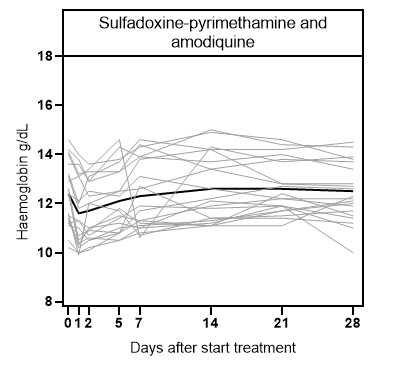

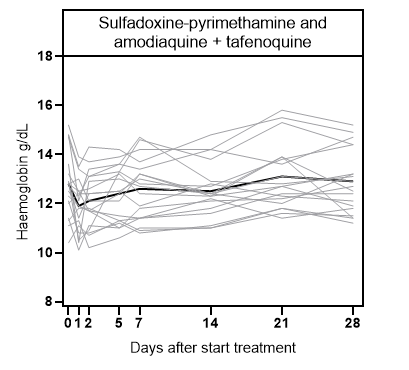

Absolute haemoglobin density is given in grams per dL (y-axis, from 8-18 g/dL) and is indicated for each participant individually with grey lines. The single black line shows the mean absolute haemoglobin density. P-values are presented in Supplementary table 7.

### Supplementary table 8. Biochemistry

| **Day of follow-up** | **Treatment arm** | **Mean ALT**  **U/L (range)** | **P-value**[^§^](https://www.thelancet.com/journals/lanmic/article/PIIS2666-5247(21)00356-6/fulltext) | **P-value**[^¶^](https://www.thelancet.com/journals/lanmic/article/PIIS2666-5247(21)00356-6/fulltext) | **Mean AST**  **U/L (range)** | **P-value**[^§^](https://www.thelancet.com/journals/lanmic/article/PIIS2666-5247(21)00356-6/fulltext) | **P-value**[^¶^](https://www.thelancet.com/journals/lanmic/article/PIIS2666-5247(21)00356-6/fulltext) | **Mean creatinine**  **mg/dL (range)** | **P-value**[^§^](https://www.thelancet.com/journals/lanmic/article/PIIS2666-5247(21)00356-6/fulltext) | **P-value**[^¶^](https://www.thelancet.com/journals/lanmic/article/PIIS2666-5247(21)00356-6/fulltext) |
| --- | --- | --- | --- | --- | --- | --- | --- | --- | --- | --- |
| Day 0 | *Overall* | 19·21 (3–105) | .. | .. | 25·09 (4–91) | .. | .. | 0·66 (0·10–1·15) | .. | .. |
|  | AL | 14·45 (3–28) | *Reference* | *Reference* | 23·20 (4–55) | *Reference* | *Reference* | 0·66 (0·42–1·11) | *Reference* | *Reference* |
|  | AL+PQ (0·25 mg/kg) | 22·90 (12–64) | *Reference* | 0·27 | 26·75 (6–53) | *Reference* | 1·00 | 0·66 (0·14–1·15) | *Reference* | 1·00 |
|  | SPAQ | 15·15 (8–29) | *Reference* | *Reference* | 21·70 (4–44) | *Reference* | *Reference* | 0·62 (0·10–1·14) | *Reference* | *Reference* |
|  | SPAQ+TQ (1·66 mg/kg) | 24·35 (5–105) | *Reference* | 0·18 | 28·70 (9–91) | *Reference* | 0·52 | 0·69 (0·24–1·14) | *Reference* | 1·00 |
| Day 2 | *Overall* | 21·43 (6–145) | .. | .. | 25·60 (3–78) | .. | .. | 0·67 (0·13–1·36) | .. | .. |
|  | AL | 15·84 (8–24) | 0·37 | *Reference* | 26·47 (17–63) | 0·17 | *Reference* | 0·55 (0·28–1·04) | 0·034 | *Reference* |
|  | AL+PQ (0·25 mg/kg) | 22·47 (10–56) | 0·92 | 0·91 | 26·47 (4–57) | 0·94 | 1·00 | 0·58 (0·17–1·04) | 0·28 | 1·00 |
|  | SPAQ | 19·20 (6–39) | 0·013 | *Reference* | 23·30 (3–38) | 0·49 | *Reference* | 0·76 (0·28–1·36) | 0·053 | *Reference* |
|  | SPAQ+TQ (1·66 mg/kg) | 28·32 (11–145) | 0·70 | 0·34 | 26·26 (5–78) | 0·64 | 1·00 | 0·78 (0·13–1·33) | 0·48 | 1·00 |
| Day 5 | *Overall* | 21·09 (6–91) | .. | .. | 25·38 (4–71) | .. | .. | 0·59 (0·13–1·38) | .. | .. |
|  | AL | 17·79 (6–48) | 0·25 | *Reference* | 23·58 (12–46) | 0·91 | *Reference* | 0·52 (0·13–1·00) | 0·003 | *Reference* |
|  | AL+PQ (0·25 mg/kg) | 21·26 (6–49) | 0·71 | 1·00 | 28·68 (7–54) | 0·55 | 1·00 | 0·54 (0·21–1·18) | 0·039 | 1·00 |
|  | SPAQ | 19·10 (7–48) | 0·062 | *Reference* | 22·85 (4–39) | 0·54 | *Reference* | 0·68 (0·14–1·38) | 0·48 | *Reference* |
|  | SPAQ+TQ (1·66 mg/kg) | 26·32 (8–91) | 0·84 | 0·53 | 26·53 (7–71) | 0·63 | 1·00 | 0·63 (0·13–1·17) | 0·28 | 1·00 |
| Day 7 | *Overall* | 20·06 (2–83) | .. | .. | 26·04 (4–73) | .. | .. | 0·67 (0·18–1·41) | .. | .. |
|  | AL | 15·21 (2–25) | 0·71 | *Reference* | 24·84 (6–49) | 0·53 | *Reference* | 0·56 (0·18–1·41) | 0·036 | *Reference* |
|  | AL+PQ (0·25 mg/kg) | 21·37 (3–40) | 0·72 | 0·61 | 25·68 (4–55) | 0·74 | 1·00 | 0·64 (0·24–1·05) | 0·59 | 1·00 |
|  | SPAQ | 20·20 (8–42) | 0·008 | *Reference* | 23·85 (6–44) | 0·31 | *Reference* | 0·78 (0·26–1·38) | 0·019 | *Reference* |
|  | SPAQ+TQ (1·66 mg/kg) | 23·47 (10–83) | 0·85 | 1·00 | 29·89 (13–73) | 0·89 | 1·00 | 0·68 (0·30–1·27) | 0·75 | 0·67 |
| Day 14 | *Overall* | 20·49 (7–50) | .. | .. | 24·57 (8–56) | .. | .. | 0·61 (0·17–1·45) | .. | .. |
|  | AL | 19·06 (9–49) | 0·12 | *Reference* | 25·33 (8–56) | 0·46 | *Reference* | 0·61 (0·17–1·38) | 0·55 | *Reference* |
|  | AL+PQ (0·25 mg/kg) | 21·68 (14–35) | 0·75 | 1·00 | 26·47 (11–47) | 0·92 | 1·00 | 0·58 (0·21–1·08) | 0·13 | 1·00 |
|  | SPAQ | 19·10 (8–49) | 0·036 | *Reference* | 21·75 (9–38) | 0·98 | *Reference* | 0·66 (0·28–1·45) | 0·41 | *Reference* |
|  | SPAQ+TQ (1·66 mg/kg) | 22·11 (7–50) | 0·65 | 1·00 | 24·89 (10–45) | 0·37 | 1·00 | 0·60 (0·17–0·97) | 0·21 | 1·00 |

Alanine aminotransferase (ALT), aspartate aminotransferase (AST) and creatinine were compared within treatment arms (p-value[^§^](https://www.thelancet.com/journals/lanmic/article/PIIS2666-5247(21)00356-6/fulltext)) using paired t-tests (with day 0 as reference) and between treatment arms (p-value[^¶^](https://www.thelancet.com/journals/lanmic/article/PIIS2666-5247(21)00356-6/fulltext)) using linear regression (adjusted for baseline levels).

AL = artemether-lumefantrine; AL+PQ = artemether-lumefantrine with primaquine; SPAQ = sulfadoxine-pyrimethamine plus amodiaquine; SPAQ+TQ = sulfadoxine-pyrimethamine plus amodiaquine with tafenoquine.
